# Estimating bone marrow adiposity from head MRI and identifying its genetic architecture

**DOI:** 10.1101/2022.08.19.22278950

**Authors:** Tobias Kaufmann, Pål Marius Bjørnstad, Martin Falck, Stener Nerland, Kevin O'Connell, Oleksandr Frei, Ole A. Andreassen, Lars T. Westlye, Srdjan Djurovic, Timothy Hughes

**Author notes:** Correspondence: Name: Tobias Kaufmann, PhD, Postal address: Department of Psychiatry and Psychotherapy, University of Tübingen, Calwerstraße 14, 72076 Tübingen; Correspondence: Name: Timothy Hughes, PhD, Postal address: Department of Medical Genetics, Oslo University Hospital, 166 Kirkeveien, 0407 Oslo, Norway.

## Abstract

Bone marrow adiposity changes radically through the lifespan, but this phenomenon is poorly characterised and understood in humans. Large datasets of magnetic resonance imaging (MRI) scans of the head have been collected to study the human brain, but also contain unexploited information about other organs. We developed an artificial neural network that localises calvarial bone marrow in T1-weighted MRI head scans, enabling us to study its composition in several large MRI datasets, and to model sex-dimorphic age trajectories, including the effect of menopause. We revealed high heritability in single-nucleotide polymorphism and twin data, and identified 41 genetic loci significantly associated with the trait, including six sex-specific loci. Integrating mapped genes with existing bone marrow single-cell RNA-sequencing data revealed patterns of adipogenic lineage differentiation and lipid loading. Finally, we identified significant genetic correlations with several human traits, including cognitive ability and Parkinson’s disease, which is intriguing in light of the recently discovered channels that link calvarial bone marrow to the meninges.

## Introduction

Bone marrow (BM) is a tissue located within trabecular bone which produces the cellular components of blood. Beginning in childhood, it undergoes a radical conversion from red to yellow adipose marrow (*1*) which begins in the long bones of the appendicular skeleton, but progresses more slowly in the flat bones of the axial skeleton (*2*). By middle-age, BM adipose tissue accounts for 50-70% of total BM volume in healthy humans (*3*). Men have higher bone marrow adiposity (BMA) than women, but post-menopausal women experience a rapid rise in BMA (*4*). The biological causes and consequences of this phenomenon are poorly understood. It is established that BM fat has a distinct function to white and brown adipose tissue depots (*5*, *6*), but a precise definition of that function and the evolutionary basis of the sex-dimorphism remain elusive. What is known is that BM adipocytes and bone-forming osteoblasts both result from the bifurcating differentiation of mesenchymal stem cells in the BM niche and this lies at the root of the well-established connection between BMA and osteoporosis (*7*–*9*). Crucially, BM adipocytes have also been shown to affect the hematopeoitic microenvironment (*10*).

Magnetic resonance imaging (MRI) scans of the head are primarily thought of as a powerful non-invasive tool for investigating the structure and function of the human brain, but these scans also contain a wealth of imaging information about other organs, such as BM. The calvarial bones contain a BM layer of approximately 600 cm^2^ which is several millimetres thick (*11*, *12*) and has recently been shown to be connected to the meninges by direct vascular channels through the inner table of the calvarium (*13*). These channels transport cerebrospinal fluid from the subarachnoid space to the cranial BM via a perivascular route (*14*, *15*) and myeloid blood cells travel in the opposite direction (*13*, *16*, *17*). Thus, there is a clear potential for the state of calvarial BM to directly affect the brain. Another specific feature of calvarial BM is that it converts from red to yellow much later in life than the BM of appendicular bones, thus its composition may be a useful normative marker of ageing.

The gold standard for assessing BM composition is magnetic resonance spectroscopy (MRS), but it is limited by its low spatial resolution, long acquisition time, and need for a predefined volume of interest, making it less well-suited for calvarial BM. Chemical shift imaging techniques offer higher resolution and shorter acquisition time, but such data are not routinely acquired. This has led to a growing interest in using conventional T1-weighted MRI to assess BM composition, with studies reporting robust correlations between BM fat volume and fraction measured with MRS, chemical shift imaging, and T1-weighted pulse sequences (*18*–*20*). Importantly, T1-weighted signal intensity is mainly determined by fat and water content (*20*–*23*) rendering it well-suited to assess BM composition.Large-scale neuroimaging studies routinely acquire T1-weighted MRI head scans, but chiefly focus on the brain. Thus, these datasets present a hitherto unexplored opportunity to address three unresolved aspects of BM biology: First, quantifying *in vivo* BM composition characteristics at population-scale, including age trajectories and sex-dimorphism, and its relationship to other traits such as bone mineral density (BMD). Second, estimating the heritability and genetic architecture of calvarial BM composition. Third, determining whether the composition of calvarial BM, which envelops the neocortex and has direct channels to it, has detectable effects on the brain.

We developed an artificial neural network-based method for locating calvarial BM in T1-weighted MRI head scans. Since T1-weighted MRI signal intensity is largely determined by fat content, we interpreted the averaged signal intensity as a proxy for calvarial BMA. We conducted extensive validation of the proposed measure using several publicly available MRI resources. We then applied this method to MRI head scans from the *UK Biobank* (*24*) to extract information on BMA, its spatial distribution in the calvarium and to model its main demographic determinants, specifically its relationship with age and sex, including female-specific factors. The size of the sample (n=33,042) enabled us to conduct a well-powered genome-wide analysis (GWAS) to identify gene associations with the proposed measure of BMA and, using a single-cell RNA sequencing (scRNAseq) dataset of BM mesenchymal cells, we gained insight into which genes drive differentiation into the adipogenic lineage and which drive lipid loading. Finally, we investigated the genetic overlap between BMA and twelve brain and body traits.

## Results

### Locating calvarial BM in head MRI scans

Our procedure for locating the BM and measuring its T1-weighted signal intensity involved 1. identifying the skin surface of the head, 2. identifying the calvarial part of the head using the top of the cerebellum and the accumbens as reference points, 3. for each point of the calvarial surface extracting an intensity array 25 mm into the skull (each layer is 0.5 mm thick), and 4. applying an artificial neural network to identify which layers of the intensity array correspond to BM (Figure 1). This network has 50 input nodes, 3 internal layers, and 6 output nodes (upper and lower bounds of outer table, BM, inner table). We trained the network on simulated data incorporating the natural intra- and inter-individual variation in thickness and intensity of the relevant anatomical structures (Figure 1A, S1). To obtain the BM signal intensity for an individual datapoint of the calvarium, we used the network model to estimate the location of the BM within the datapoint’s intensity array and averaged these BM intensities to get the BM intensity for that datapoint. Then, we averaged these datapoint intensities across the calvarium to produce the global BMA measure for the scan.

**Figure 1:**
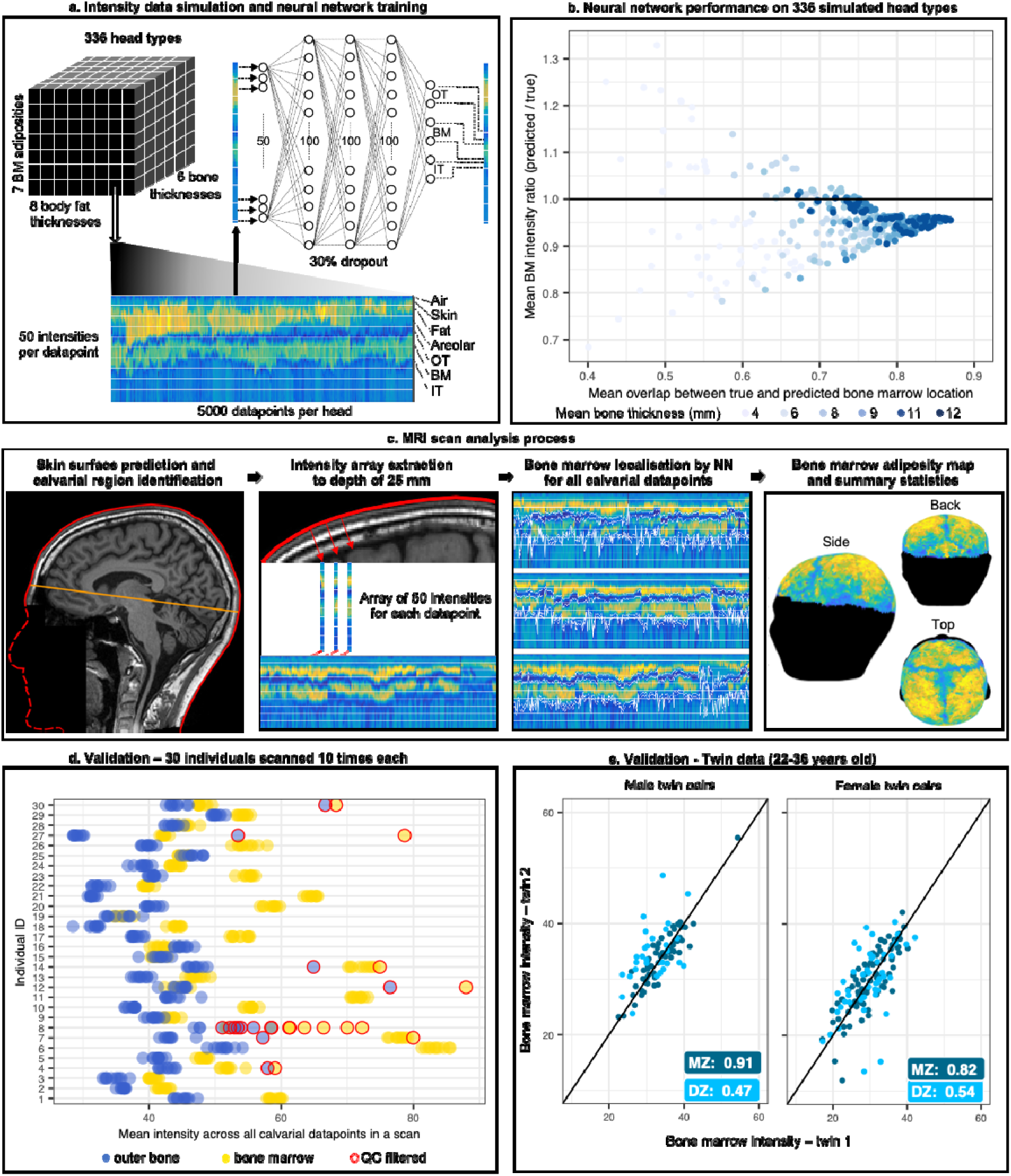
Overview of the artificial neural network training, application, and validation. **a**. The data used for training the network were generated by simulating 336 different head types (with an example simulation for one head type provided here), where a head type is defined by mean bone thickness, mean BM intensity and mean body fat thickness. There are 5000 datapoints per head each with 50 intensities per datapoint. OT: outer table, IT: inner table, BM: bone marrow. **b.** We evaluated the performance of the neural network using two metrics: 1. the overlap between the predicted and the true BM location (defined by the simulation), and 2. the ratio between the predicted signal intensity of the BM and the true intensity of the BM. **c.** Illustration of the process of extracting the intensity arrays from the MRI scan, locating the BM, and computing its intensity. **d.** The algorithm was validated on a real dataset consisting of 30 individuals that were scanned 10 times each at 3-day intervals. For each scan, we computed the intensity of the outer bone (blue) and of the BM (yellow) which displayed high concordance across scans of the same individuals. The quality control measures correctly identified scans not producing reliable measures (13 of 300, 11 of which fail due to too few datapoints). **e.** Application of the algorithm to a cohort of monozygotic and same-sex dizygotic twins. Correlation in BMA between twin pairs in coloured boxes.

### Procedure validation on simulated data

We evaluated the performance of the neural network on simulated data using two metrics (Figure 1B): 1. the overlap between the predicted and the true BM location, and 2. the ratio between the predicted intensity of the BM and the true intensity of the BM. The median overlap between the predicted BM and the true BM in the simulated dataset was 0.76 with the lowest value (0.4) obtained in calvaria with a thin bone and the highest value (0.88) obtained in calvaria with thick bone. Poor overlap (below 0.7) was almost only observed in the thinnest bone and is explained by the fact that when the BM part of the bone is only a few layers thick (1 layer = 0.5 mm), an error by one layer will inevitably lead to a substantial fall in overlap. However, this did not result in a corresponding fall in the ratio of the predicted intensity of BM to its true intensity, because the typical BM intensity was only marginally higher than the neighbouring bone intensity. This property of the typical relative intensities of these anatomic structures also explained why the intensity ratio at high overlap is not centred on 1: any misidentification of cortical bone as BM, will typically result in an underestimate of true BM intensity (Figure S2). The neural network performed well and intensity ratios were in the range 0.9-1.1 for the vast majority of head types (Figure S3). It was only for heads with thin bone and either very high or very low BM intensity that we observed intensity ratios below or above this range (Figure S3B). Variation in subcutaneous fat thickness had no systematic impact on the performance (Figure S3C).

### Procedure validation on real data and heritability estimate

The procedure also performed well on real data. We first tested a dataset from the *Consortium for Reliability and Reproducibility* (*25*) consisting of 30 individuals that were scanned 10 times each at 3-day intervals. The scans passing quality control displayed mean intensity of the outer table and BM at the levels that we would expect (Figure 1D): very low variation between scans of the same individual, outer table intensities centred on similar values with low variation across individuals, BM intensity always higher than outer table intensity, and high variation of BM intensity between individuals, but little within individuals. These scans were also inspected visually to verify that structure boundaries were correctly identified. Figure S4 provides examples of network performance in data of individuals with different levels of subcutaneous fat thickness, BMA, and bone thickness. Furthermore, we performed several assessments of the impact of different intensity normalization approaches on the obtained BMA estimates and compared estimates based on T1-weighted images to estimates obtained from T1 relaxation maps derived from a quantitative MRI pulse sequence (MP2RAGE). The BMA measure derived from T1-weighted images significantly correlated with BMA derived from the quantitative T1 relaxation maps and was not prone to differences in standard image normalization procedures (Figure S9-S12), supporting the validity of the proposed approach.

The monozygotic (MZ) twin data from the *Human Connectome Project* cohort (*26*) (Figure 1E) provided a second validation of the procedure through the high concordance in mean BM signal intensity between MZ twins, 0.91 in male pairs and 0.82 in female pairs. Further, using the data on the same-sex MZ and DZ twin pairs, we showed that BMA has an estimated broad-sense heritability of 91% in males and 60% in females (Table S1). We estimated a strong influence of environmental factors in females, one of which is highly likely to be estrogen levels as these vary widely between women of child-bearing age, with particularly large changes during pregnancy when estrogen levels are orders of magnitude higher, and the common use of estrogen-based contraceptives.

### The calvarial BMA phenotype

In the 16,140 male and 16,902 female white British individuals from the *UK Biobank* for which head imaging data is available, we observed that calvarial BMA had strongly sex-dimorphic features. Males had a higher mean BMA than females (Figure 2A) and showed no increase at the group level in BMA levels in the age range covered by the biobank sample (45 to 82 years) implying that the rise in BMA must have occurred earlier in life (Figure 2B). In females the BMA level was highly dynamic in this age range increasing by 50% in the same age interval and thereby reaching male levels at age 70. Hormone replacement therapy (HRT) was used at some point in life by 38% of the women in the study and this therapeutic intervention slowed the rise in BMA, resulting in average BMA levels for treated women that did not exceed the level of males at age 80.

**Figure 2:**
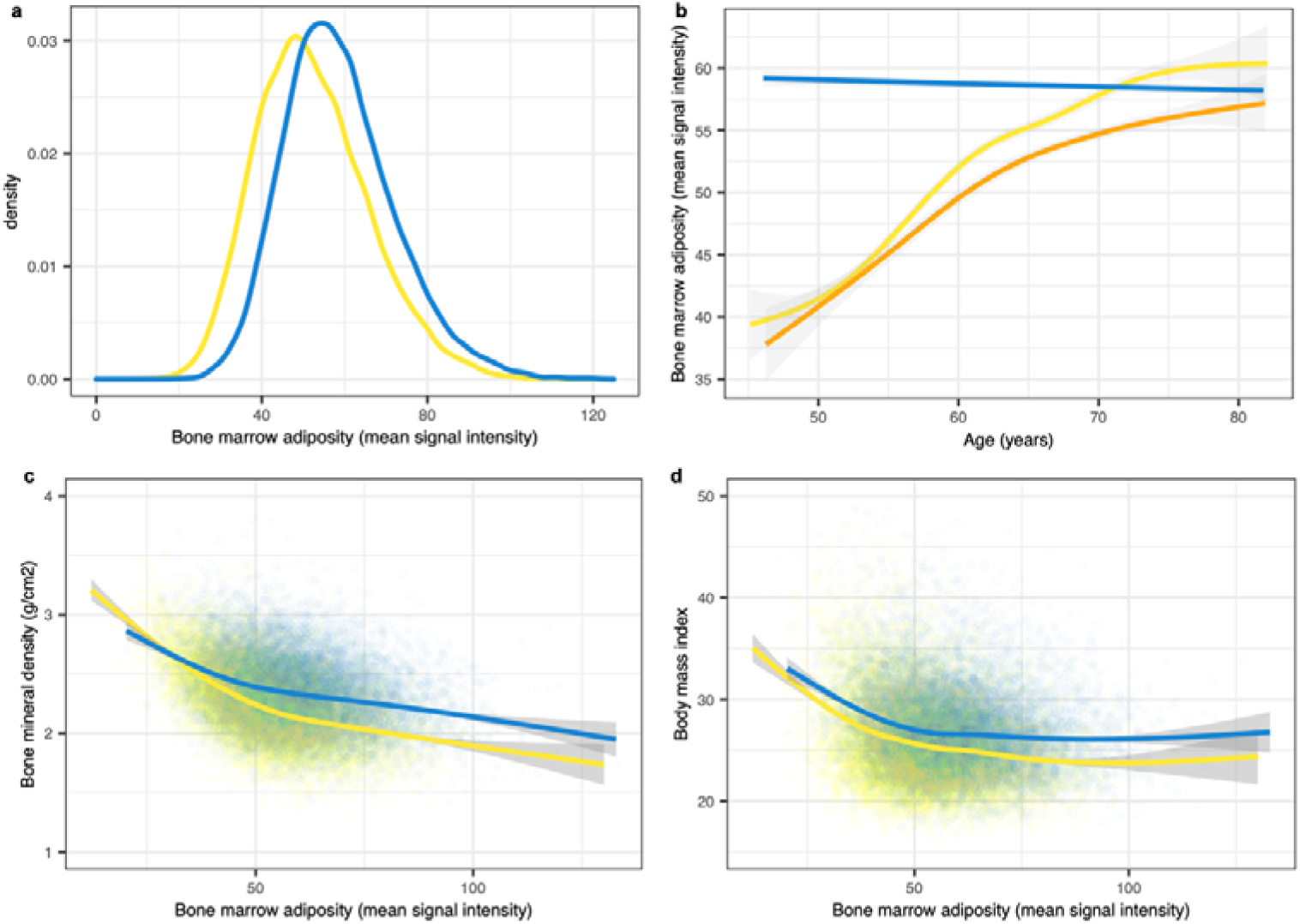
The calvarial BMA phenotype. **a.** BM intensity distribution in males (blue) and females (yellow) **b.** BMA age trajectories for males and females. Males in blue and females in yellow (never HRT) and orange (ever HRT). The effects of age and HRT on female BMA levels were both statistically significant (p-value < 2.0E-16) **c.** Sex-dimorphic relationship between BMA and calvarial bone mineral density. **d.** Relationship between BMA and BMI. Fitted lines in B, C, and D were computed using a general additive model and shading represents 95% confidence intervals.

BM adipocytes and osteoblasts differentiate from the same mesenchymal progenitor cells and this would appear to drive the known negative correlation between BMA and bone mineral density (BMD). We replicated this effect in our data (Figure 2C – 14,262 males and 14,831 females). This relationship is also dimorphic with females displaying a lower level of calvarial BMD for a given level of BMA. Further, the size of the female BMD deficit increases as BMA levels rise (a similar pattern is observed with BMI in Figure 2D). We quantify the determinants of calvarial BMD by linear regression (Table S2). These regressions confirm the known effects of age and sex, and that the effect of BMA on BMD is about twice as large in females relative to males. Our regression results suggest that most of the effect of HRT on BMD is mediated by its effect on BMA (Table S2) since: 1. the statistical significance of HRT as an explanatory variable for BMD disappears when BMA is included as an explanatory variable in the regression and 2. the R^2^ of the regression increases by a factor of 4 to 30%.

### Genetic architecture

The strong twin-based heritability of BMA emphasised the importance of a genome-wide association study (GWAS) of the trait. We used the *UK Biobank* cohort (Table S3) and performed sex-specific covariate regression prior to separate discovery GWASs on males (n=16,140) and females (n=16,902). We found the male and female GWASs to have low genomic inflation (Figure 3A and Table S4) and to be significantly genetically correlated (Rg=.94, P=6e-27, Figure 3B), allowing us to combine these samples to perform a joint discovery GWAS across males and females (n=33,042). Further, we performed an independent replication GWAS (n=4,958) on non-white British males and females.

**Figure 3:**
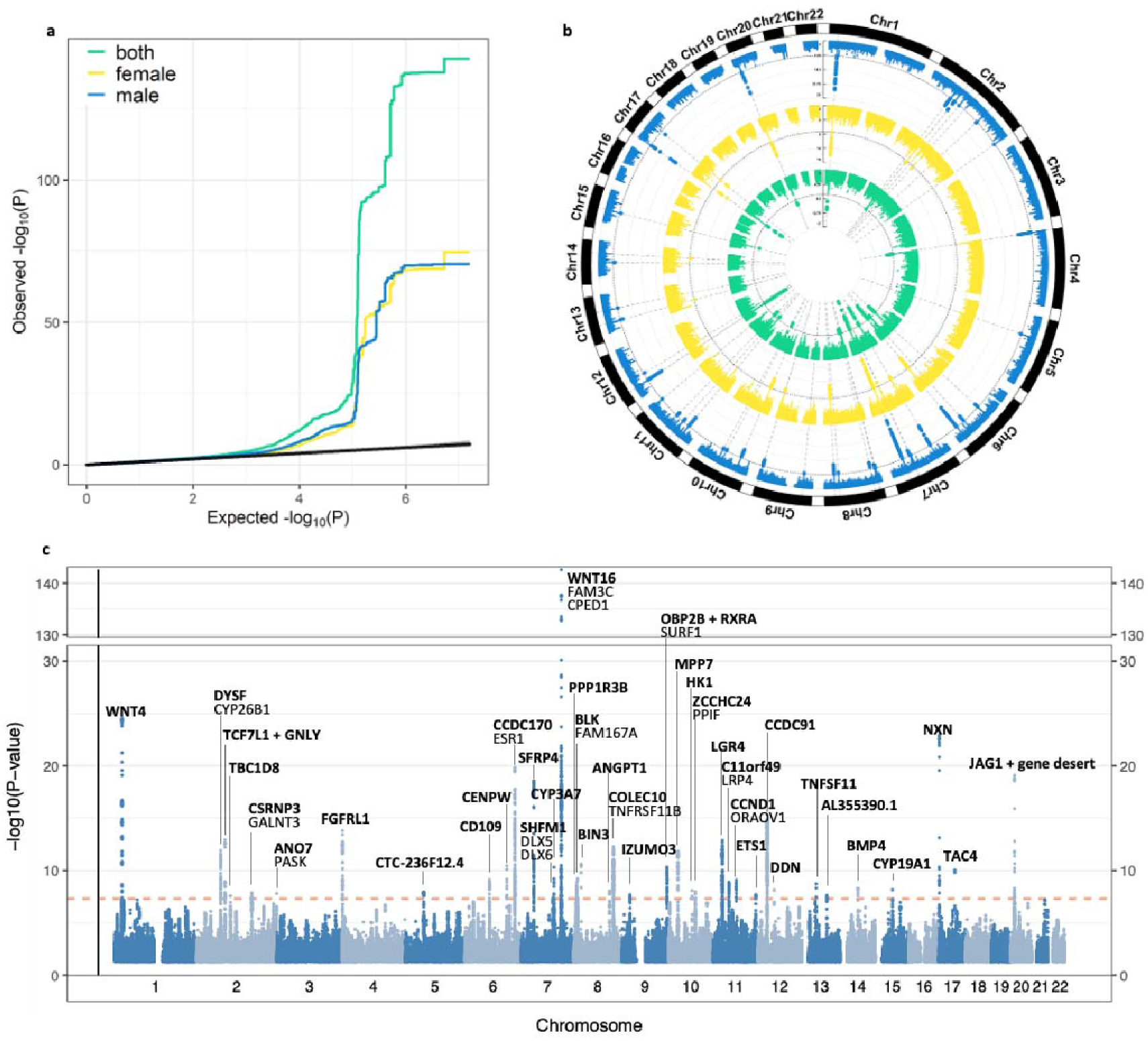
BMA genome-wide significant loci. **a.** QQ-plot of p-values for the male, female and joint discovery GWAS **b.** Comparison of male (outer), female (middle), and joint (inner) discovery GWAS significant loci. **c**. Manhattan plot of the 41 genome-wide significant loci in the joint discovery GWAS named after gene closest to the top lead SNP (bold). Other genes in each locus for which genome-wide significant SNPs were eQTLs, are listed below the closest gene. Three loci were closely adjacent to another more significant locus and are marked “+”. SNPs with p < 0.05 not plotted.

SNP-based heritability (h^2^_SNP_) estimated using linkage disequilibrium score regression confirmed significant heritability of the joint discovery GWAS, (h^2^_SNP_ = 31.5%) (Table S4). We identified 168 genome-wide significant independent SNPs (P<5e-8), corresponding to 41 independent genomic loci (Table S5). The replication GWAS also displayed significant heritability (h^2^_SNP_ = 28.6%). Out of the 168 significant discovery SNPs, 62% replicated at P<.05, and 39% of the 41 lead SNPs replicated at P<.05 (Table S6). One locus replicated at genome-wide significance (P<5e-8). Furthermore, 92.7% of the lead SNPs of the discovery sample showed same effect direction in the replication sample (Table S5).

Figure 3C depicts genes mapped from the 41 loci associated with the calvarial BMA. The most significant top lead SNP was located in the WNT16 gene (P=2.6e-143), which is also the top hit in BMD GWAS (*27*, *28*) (Figure S12). The ESR1 locus displayed a notable sex-dimorphic pattern (Figure S13). In females, the top lead SNP overlapped the ESR1 gene, but another lead SNP overlapped the neighbouring CCDC170 gene. In males, the signal in ESR1 was absent, but a genome-wide significant signal was located in the CCDC170 gene, suggesting that the ESR1 gene might not be the only source of an association signal in this locus.

A comparison of the male and female effect sizes of the top lead SNPs of each locus revealed 6 loci in which there is a more than two-fold difference between the sexes (loci 10, 18, 26, 30, 32, 37 in Table S5). In all cases, it is the male effect size that is larger. Further, each locus is genome-wide significant in males, but not in females, thus suggesting that these loci may be genetic drivers of the sex-dimorphism.

### Gene expression in BM mesenchymal stem cell differentiation

Mesenchymal stem cells of the BM niche commit to either the adipogenic or the osteogenic lineage (Figure 4A) and both the number committing to the adipogenic lineage and their level of lipid-loading influences the total level of BMA. This aspect of BM biology is shared between humans and mice (*29*), so we made use of an existing mouse scRNAseq dataset of BM mesenchymal lineage cells (*30*) to study variation in the expression of BMA-associated genes as cells differentiate (Figure 4B). For any given gene, the percent of cells expressing a gene was relatively stable across different cell types. However, the mean scaled expression varied considerably between differentiation stages and the clustering of expression profiles revealed that most genes fall into one of the two largest groups of profiles: either genes with their peak of expression at the lineage commitment stage (LMP and LCP) or within the adipogenic lineage (MALP). It was also noteworthy that the genes peaking in MALPs mostly showed low expression prior to that stage. This differed from the ramping up observed in mesenchymal progenitors and LCPs. Only one of the closest genes saw their peak of expression in osteoblasts, but it was unexpected to observe that 6 of the closest genes see a strong peak in osteocytes. It remains to be investigated whether the closest gene might not be the source of the association or whether gene expression in osteocytes may have an influence on BMA.

**Figure 4:**
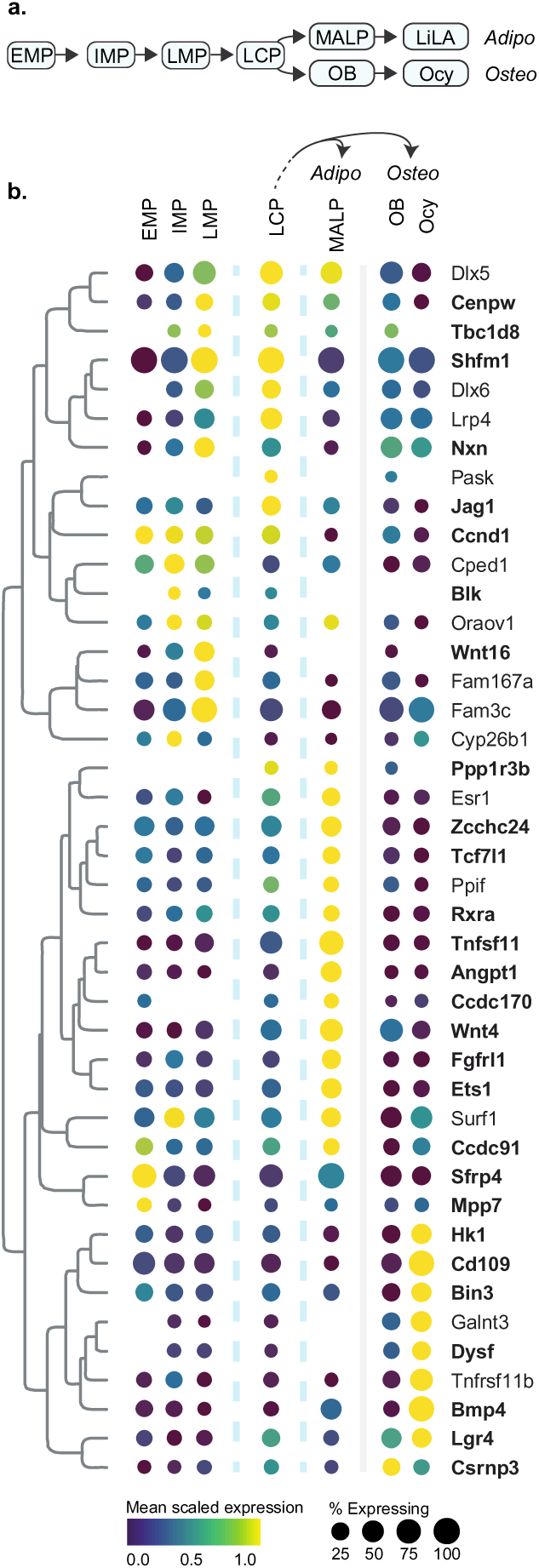
Gene expression in mesenchymal lineage cells of mouse BM. **a.** The differentiation of BM mesenchymal cells. Early (EMP), intermediate (IMP), and late mesenchymal progenitors (LMP). Lineage committed progenitors (LCP). Marrow adipogenic lineage precursor (MALP). Lipid-laden Adipocyte (LiLA). Osteoblast (OB). Osteocyte (Ocy). **b**. A clustering of the gene expression profiles across cell types. Gene names are for mouse genes (these broadly match human gene names). Of the 40 genes which are closest to lead SNP for each locus, 28 had expression in the scRNAseq data (bold). LiLA expression data was not available in this dataset.

### Genetic correlation and overlap

We investigated genetic correlation and overlap with 13 traits. We selected body traits (BMD, BMI, waist-to-hip ratio, systolic and diastolic blood pressure, type-2 diabetes, coronary artery disease) that have a logical connection to BMA given the mesenchymal stem cells origin of BM adipocytes and their role in bone, fat, and vasculature (*30*). For the brain, we selected traits reflecting cognition (general cognitive ability and educational attainment) and disorders that are prevalent in adulthood (insomnia, multiple sclerosis, Parkinson’s disease, Alzheimer’s disease) since it is primarily in adulthood that the adiposity of calvarial BM experiences a substantial change (Figure 2B).

We identified strong negative genetic correlations between BMA and both BMD and BMI, and more subtle global genetic correlations between BMA and Parkinson’s disease, general cognitive ability, and educational attainment (Table 1). However, global genetic correlations lack the ability to assess genetic overlap in the presence of mixed effect directions. So, to better understand the genetic overlap between BMA and other traits of interest, we also applied MiXeR (*31*). MiXeR uses trait polygenicity, defined as the number of trait-influencing variants required to explain 90% of SNP-based heritability, to model the number of shared trait-influencing-variants between two traits as well as the correlation within this shared component. MiXeR estimates that 99% of BMA trait-influencing variants are shared with BMD (497 of 500 variants), and that the correlation of this shared component is -0.95, indicating high genetic overlap with discordant effect. The proportion of BMA trait-influencing variants also estimated to influence general cognitive ability (0.57) and educational attainment (0.67) was lower than observed for BMD, but the correlations within their shared components were similarly large (COG = 0.93, EDU = 0.90). These patterns of high overlap and high effect correlation within this overlap are suggestive of vertical pleiotropy i.e. a molecular mechanism influencing one trait, that in turn influences a second trait, such that most of the variants driving the first trait either have the same or the opposite direction of effect on the second trait.

**Table 1:**
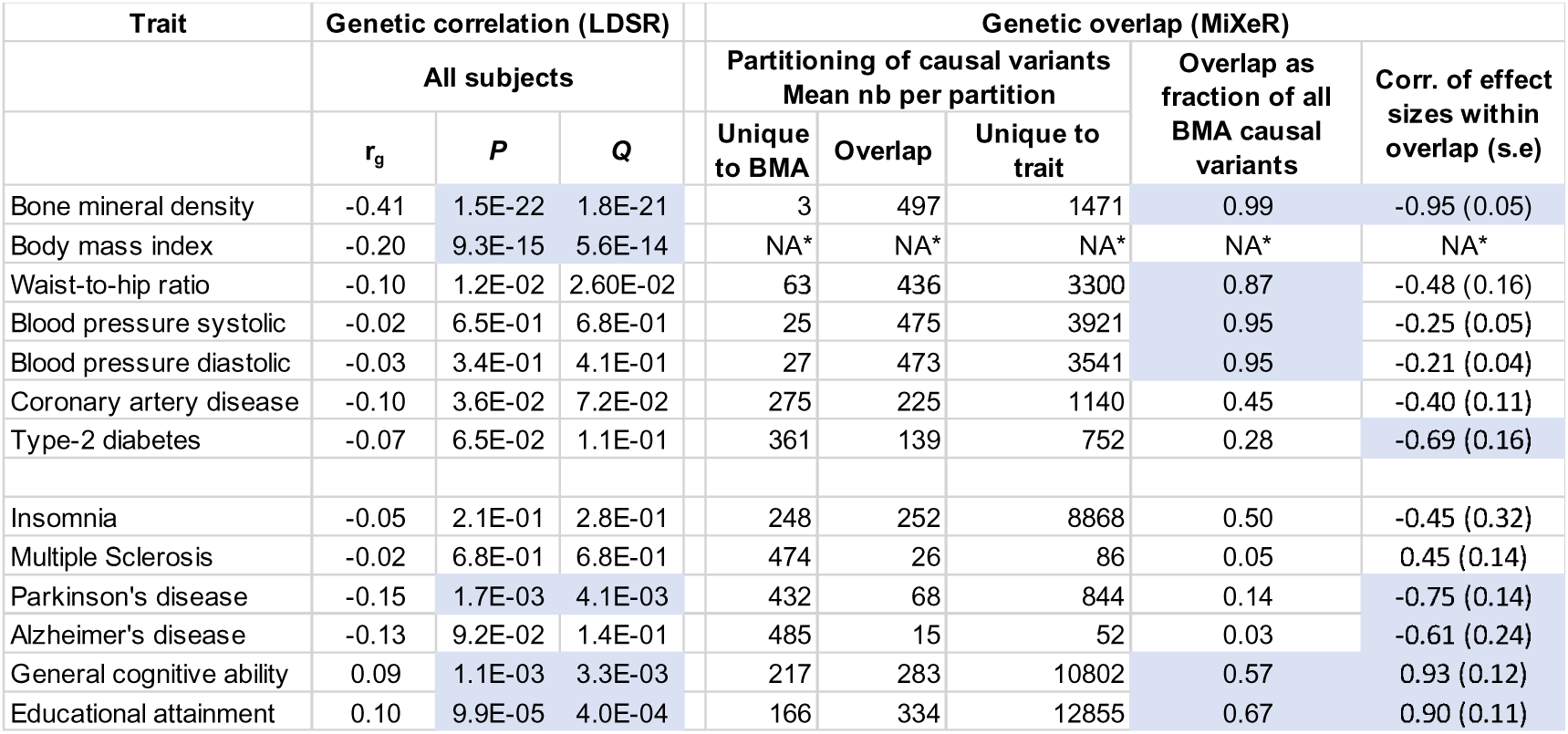
Genetic correlation and overlap of BMA with other traits. r_g_: genetic correlation. *Q*-values are FDR corrected *P*-values. Genetic overlap: number of causal variants that overlap (i.e. are shared) between BMA and the second trait. Total number of estimated causal variants for a trait (i.e. explaining 90% of SNP heritability) is the sum of those unique to trait and overlap (large total is indicative of high polygenicity). NA*: The publicly available summary statistics of the BMI GWAS contained only 3.3M SNPs, it was thus not possible to apply the MiXeR method to this dataset. Sex-specific genetic correlations, standard errors for causal variant partitioning, and Akaike Information criterion values (Table S9). Blue highlighting for: genetic correlation p < 0.01, overlap fraction > 0.5, correlation of effect sizes within overlapping > 0.5 or < -0.5.

In Parkinson’s Disease, where we also estimated statistically significant genetic correlation with BMA, MiXeR estimates a strong negative correlation of effect sizes, but this is within a much lower percentage of BMA trait-associated variants (14%). Similarly, BMA displayed a high correlation of effect sizes (-0.69) with Type-2 diabetes but within a moderate overlap, and a low correlation of effect sizes with both systolic and diastolic blood pressure, but with an overlap of 95%. These patterns are more indicative of horizontal pleiotropy (i.e. molecular mechanisms influencing multiple traits) than vertical pleiotropy.

## Discussion

The development of a method for locating BM in MRI head scans enabled us to characterize calvarial BMA in several datasets and to quantify, *in vivo* and at population-scale, BMA’s age trajectory, sex-dimorphism, and relationship to BMD. We focused on BM adiposity rather than volume, as this trait was considered more likely to be influencing other traits of interest, such as the blood (hematopoiesis), bone (osteoporosis), and brain health (through transcortical channels). Indeed, through a well-powered GWAS, we identified the main genetic drivers of this phenotype and we observed significant genetic correlation and overlap with several traits, including Parkinson’s disease and general cognitive ability.

### Estimating BMA in MRI head scans

BMA can be quantified in whole-body scans (*32*), but calvarial BM has a number of features which make it particularly well-suited to the study of BMA. First, conversion to yellow BM is slower in the flat bones (*33*), such that sex-dimorphic trajectories are observable in middle-aged cohorts. Second, there are many large collections of T1-weighted MRI scans from which calvarial BMA can potentially be extracted and, although T1-weighted pulse sequences are not specifically designed for BMA quantification (*34*), this enables analyses that have not previously been possible, such as GWAS and accurate estimation of lifespan sex differences. Finally, it has a simple “sandwiched” structure that we were able to model and then generate simulated calvaria which we use as training data for an artificial neural network. Our validation procedures demonstrate that this automated BM location procedure delivers good performance on real data, even though it was trained on simulated data and analyses each datapoint independently of all others. A more sophisticated model (*35*, *36*), trained directly on annotated real data, may raise performance further, for example by improving the prediction of the lower bound of the BM. However, this would require a substantial manual annotation effort and would face the difficult challenge of distinguishing low-adiposity BM from calvarial bone. Both these issues are circumvented by our approach.

### The calvarial BMA and BMD phenotypes

The sex-dimorphic nature of BM adiposity is qualitatively well-established (*4*), but has not previously been quantified in a large human cohort. Males have a high and stable average level of BMA over the covered age range (45 to 82 years of age), indicating that the dynamic male phase occurs earlier in life. The youngest females in our sample have a low level which ramps at a constant rate from the beginning to the end of the range with HRT significantly reducing the rate of increase. Interestingly, however, by the age of 70 (for women not using HRT), female BMA has risen to the same level as males. It appears highly likely that the menopause and the accompanying drop in natural estrogen levels is part of what drives the rise in BMA levels and that estrogen treatment can delay the rise. We emphasise that these trajectories are for calvarial BM and are most probably different in profile and levels to other BM, since it is known that axial flat bones convert later and perhaps less completely than the appendicular long bones. However, the nature of the sex-dimorphism and the effect of estrogen is likely shared with other BM.

Women at risk of osteoporosis are often prescribed HRT in order to improve BMD. Our regression results (Table S2) indicate that the effect of HRT on BMD is mostly mediated by its effect on BMA. Further, we suspect that the effect of BMA on BMD may be underestimated given that the prescription of HRT is biased towards women with low BMD and high BMA. Future studies, including information on why HRT was prescribed and for how long, may use our BMA quantification method to further detail this important relationship.

### BMA heritability estimates

Our findings showed that male SNP heritability (43%) is half the broad-sense heritability (91%). In females, SNP heritability is only 23% which is low relative to men and relative to the broad-sense heritability of 60%. Since the vast majority of women in the *UK Biobank* sample have probably not reached their ultimate BMA levels, we suspect that the genetic effects on the phenotype may be underestimated in this female GWAS sample. Despite these features of the sample and its relatively modest size, we do observe good genetic correlation between the sexes of the discovery sample (Figure 3b) and cross-ethnic replication in a limited sample of 4,958 individuals. Based on these GWAS results, MiXeR estimates that BMA has approximately 500 causal variants, indicating low polygenicity relative to a typical polygenic trait such as BMD, which we estimated to have approximately two thousand causal variants.

The high broad-sense heritability estimates indicate that environmental effects on BMA are limited relative to genetics. However, a limitation of our broad-sense heritability estimates is that the age range of the twin dataset is much younger than the *UK Biobank* dataset from which we compute SNP heritability. This is particularly relevant for females, first because fluctuations in estrogen levels are substantial during child-bearing age due to pregnancy and contraception, and second because females only reach their ultimate BMA level between the ages of 70 and 80 (the upper end of the age range). Both the broad-sense and SNP heritability estimates should therefore be interpreted with caution for females.

### Genetic correlation and overlap with other traits

We selected 7 body and 6 brain traits for which we computed genetic correlation, and obtained significant results for BMD, BMI, Parkinson’s disease, general cognitive ability and educational attainment. Genetic correlation has two limitations. First, it tends to underestimate the true correlation in the case of mixed effects and, second, it provides no indication of whether the relationship between the traits is driven by horizontal or by vertical pleiotropy. MiXeR estimates the causal variants for each trait and thereby can estimate the overlapping fraction as well as the correlation of effect within this set. A high overlap of BMA causal variants with those of the other trait and a high correlation of effects within this set is suggestive of causal effect (vertical pleiotropy). Only 3 of the 12 traits displayed a clear combination of such features. One of these is BMD, which can be considered as a positive control since the BMA and BMD traits are known to be negatively correlated and the bifurcating differentiation of mesenchymal stem cells provides a causal cellular mechanism for this relationship between traits. The other two traits are educational attainment and general cognitive ability. Since these are two highly correlated traits (*37*), a high overlap and correlation of genetic effects for both traits with BMA may be consistent with the hypothesis that BMA could have a causal effect on cognition (Table 1).

### GWAS results and differentiation of mesenchymal progenitors

Previous studies of mesenchymal cell differentiation in animal and cell models have pointed towards the Wnt/β-catenin signalling pathway being an important determinant of BMA (*29*). Our large-scale phenotyping of BMA in a human cohort enabled us to perform a hypothesis-free quantification of its genetic architecture which confirmed the strong effect of genes related to this pathway, including WNT16 (*38*), WNT4 (*39*), NXN (*40*). Further, we found associations in genes (BMP4 (*41*, *42*), DLX5 (*9*, *43*), LGR4 (*44*)) or close paralogs of genes (LRP4/LRP5(*45*), SFRP4/SFRP1 (*46*)) that were known to specifically influence adiposity. We also discovered and quantified the effect of many interesting novel loci, such as FGFRL1 which has a pleiotropic role in hypertension resistance and bone growth in giraffes (*47*). The strong effects of the CCDC170/ESR1 and WNT16 loci (Table S5) were not unexpected given previous results from the BMD GWAS (*28*), but the pattern of the genetic signal illustrates the importance of fine mapping for these and other loci (Figures S9-10).

Recent scRNAseq studies of the BM niche have provided a comprehensive insight into BM cell types and the changes in gene expression during differentiation (*48*–*50*). Zhong et al. focused specifically on mesenchymal lineage cells and produced a detailed transcriptomic profile of progenitor maturation and bifurcating differentiation into adipogenic and osteogenic lineages (*30*). Interestingly, they identify a new non-proliferative cell type in the adipogenic lineage that does not contain lipid droplets (MALP: marrow adipogenic lineage precursors). In our clustering of GWAS genes according to their scRNAseq transcriptomic profiles, we find 17 genes which ramp expression in mesenchymal progenitors with the peak of expression occurring in LMP or LCP, and 16 genes with very low expression in mesenchymal progenitors and a distinct peak at the MALP stage. Since BMA is primarily determined by two potentially asynchronous processes, the number of adipocytes and the level of lipid-loading, this suggests that the first set (including WNT16) controls the bifurcation between the two main lineages, whilst the second set (including ESR1) controls MALPs further differentiation into lipid-laden adipocytes.

MALPs outnumber lipid-laden adipocytes by orders of magnitude in young mice and exist as pericytes and stromal cells that form a ubiquitous 3D network inside the marrow cavity (*30*). Evidence points to this network playing pivotal roles in maintaining marrow vasculature (*30*), suppressing osteogenic differentiation (*51*), and probably also influencing hematopoiesis (*10*). We tentatively speculate that calvarial MALPs may be involved in sensing perivascular flows of CSF from the meninges to the BM and in influencing the BM’s hematopoietic response, and that this might be the basis of the observed genetic overlap between BMA and some cerebral traits. However, more conventional anatomical pathways may also be relevant, as the diploë and inner table communicate with meningeal and dural venous systems through diploic veins.

In addition to these recently described channels, classical cranial venous anatomy provides other potential interfaces between calvarial marrow and the intracranial compartment, including diploic and emissary venous connections with meningeal veins and dural venous sinuses.

### Limitations and future validations

Several previous studies have used T1-weighted MRI to study BM composition (*52*–*54*). Validation studies have reported correlations of 0.99 with BM fat volume and of 0.71 with BM fat fraction measured with the IDEAL technique in the tibia of children (*18*). Similarly, BM fat volume estimated with T1-weighted images have shown strong correlations with MRS (r=0.88) and the Dixon method (r=0.79) for vertebrae and with the Dixon method for femoral neck (r=0.86) in menopausal women (*19*). A score based on T1-weighted signal intensities in the vertebrae was found to be a promising marker for osteoporosis and correlated with BMD measured with DEXA (r=-0.68) (*55*). While most prior studies have focused on non-cranial BM, one study found that T1-weighted signal intensity in the calvarium was a sensitive (93%) and specific (86%) marker for infiltrative processes including lymphoma, leukaemia and chronic anaemia known to affect BM composition (*20*).

Whereas prior studies have relied on manual grading and selection of segmentation thresholds and regions of interest, our approach automatically locates calvarial BM. This improves repeatability and allows for deployment in large datasets such as the UK Biobank. Although the main determinant of the T1-weighted signal intensity is fat, making it well-suited to assess BMA, weighted MRI pulse sequences present notable challenges. Importantly, T1-weighted signal intensity is influenced by scanner-related variation (*56*).

To ensure between-subject comparability, we used the intensity normalised nu.mgz volume output by FreeSurfer. We validated this approach through comparison with well-established intensity normalization methods: Kernel Density Estimation (KDE), WhiteStripe (WS), Gaussian Mixture Model (GMM), Fuzzy C-Means (FCM), and Z-score normalization (ZS). We found the highest test-retest reliability with our approach (Figure S9), and, together with KDE (based on reference signal intensity in WM), the highest correlation with quantitative T1 relaxation maps (Figure S10). Together with the significant correlations with BMD (Figure S11), as measured with DEXA, and significant effects of osteoporosis status (Figure S12), these results demonstrate the biological relevance of the proposed measure of BM composition.

Nevertheless, we recognise the need for further validation to disentangle the relative contributions of adiposity and other features of BM composition. As such, caution is still warranted on the biological interpretation of measures based on T1-weighted images, and future sensitivity and robustness analyses are needed. Given the plausible biological findings in our study (sex-dimorphic age trajectories mapping known BMA biology, discovery of genes previously identified through cellular experiments as critical to adipocyte biology), we are confident that the benefits of exploiting the existing large samples of T1-weighted images outweighs the limitations. Indeed, future studies can build on our developments to further validate the proposed measure of bone marrow composition and to study its effect on bone, blood, and brain.

## Materials and Methods

### Experimental design

This study had several sequential subgoals: 1. obtain data on bone marrow composition from head MRI scans, 2. determine the sex-dimorphic age trajectories, 3. identify the genes associated with this trait, 4. determine which of these genes control the bifurcating (osteogenic/adipogenic) differentiation and which control lipid loading and 5. investigate whether the bone marrow composition trait has genetic overlap with other related body or brain traits. Each of these study components requires specific methodology and several data sources are used. These are described separately in the following subsections.

### Cohorts

#### Hangzhou Normal University (HNU) data as part of the Consortium for Reliability and Reproducibility (*25*)

We accessed the openly available test-retest reliability dataset from http://fcon_1000.projects.nitrc.org. In this study, T1 MRI was acquired on a 3T GE Discovery MR750 scanner using an 8-channel head coil with a repetition time of 8.06ms, 8° flip angle and 250 mm field of view at a resolution of 1x1x1 mm. We included all available data from this sample, specifically data from 30 healthy adults (50% female) aged 20-30 years (mean: 24.4, sd: 2.4 years) that were scanned ten times across one month, one scan every three days.

#### MICA-MICs data (*57*)

We accessed the openly available multimodal dataset from https://portal.conp.ca/dataset?id=projects/mica-mics. In this study, MRI data was acquired on a 3 T Siemens Magnetom Prisma-Fit equipped with a 64-channel head coil. T1 MRI was acquired with a 3D magnetization-prepared rapid gradient-echo sequence (MPRAGE; 0.8 mm isotropic voxels, matrix = 320 × 320, 224 sagittal slices, TR = 2300 ms, TE = 3.14 ms, TI = 900 ms, flip angle = 9°, iPAT = 2, partial Fourier = 6/8). Quantitative T1 MRI was acquired with a 3D-MP2RAGE sequence (0.8 mm isotropic voxels, 240 sagittal slices, TR = 5000 ms, TE = 2.9 ms, TI 1 = ld0 ms, T1 2 = 2830 ms, flip angle 1 = 4°, flip angle 2 = 5°, iPAT = 3, bandwidth = 270 Hz/px, echo spacing = 7.2 ms, partial Fourier = 6/8).

#### Human Connectome Project data (*58*)

We accessed openly available data from https://db.humanconnectome.org. In this study, T1 MRI was acquired on a customized 3T Siemens Skyra scanner with a repetition time of 2400ms, 8° flip angle, 224mm field of view and a voxel size of 0.7mm. We included data from 430 twins from this sample, including 174 males (60 dizygotic and 114 monozygotic, age range 22 to 34) and 256 females (96 dizygotic and 160 monozygotic, age range 26 to 36).

#### UK Biobank (*24*)

We accessed data from the UK Biobank study resource (access code 27412; https://www.ukbiobank.ac.uk). In this study, T1 MRI data was acquired on three identical 3T Siemens Skyra scanners with a repetition time of 2000ms, 8° flip angle, 256mm field of view and a resolution of 1x1x1mm. We split the available sample into a discovery set comprising 33,042 white British individuals (51% females) aged 45-82 years (mean 64.9, sd 7.5 years), and an independent replication set comprising all other individuals (N=4958, 52% female, age mean 62.9, sd 7.7, range 45-81 years). We also obtained head bone mineral density measurements produced by a DXA system (code 23226-2.0), body mass index (code 21001-2.0), and data on whether women ever used hormone-replacement therapy (code 2814-2.0). BMD data was only available for 29,093 of the samples for which MRI head scans were available.

### Ethical approvals

The UK Biobank was approved by the National Health Service National Research Ethics Service (ref. 11/NW/0382). The Human Connectome Project was approved by the Washington University institutional review board. Hangzhou Normal University study protocol and data sharing was approved by its local ethics committee.

### FreeSurfer preprocessing and extraction of datapoint intensities

We processed all T1 MRI data using the standard recon-all pipeline in FreeSurfer 5.3.0. T1 intensities were extracted from the nu image, which is corrected for intensity non-uniformities and intensity normalized via scaling. Next, we fed the intensity-normalized volumes derived via recon-all into the FreeSurfer mri_watershed program to delineate the surfaces of the brain, skull and skin. We performed a stepwise extraction of intensities from layers following the shape of the outer skin, walking inward into the skull in steps of 0.5mm up to 25mm inward which produces an intensity array of length 50 for each datapoint. Since watershed delineates the skin around the entire head, we disregarded parts that are not directly on top of the cortex. For this, we identified the coordinates of the upper part of the cerebellum and the accumbens from the individual level FreeSurfer aseg parcellation and defined a plane in 3D space that intersected these two points, disregarding all data points in coordinates below the plane. The resulting stream of layerwise intensities therefore comprised data on top of the cortex and was fed one-by-one into an artificial neural network model to identify and define the borders of the bone marrow (BM) for that datapoint. See supplementary materials for more detail on this workflow, including software versions.

### Intensity array data simulation

In order to train the neural network model, we generated a large synthetic dataset of intensity arrays, with known boundaries between anatomical structures, by simulating the thickness and intensity of the different structures located between the outer skin and the sub-arachnoid space. The simulation incorporated the following real-world complexities:

1. Different anatomical architectures (skin, subcutaneous fat, aponeurosis, outer table, BM, inner table, dura mater, arachnoid space), including when a structure is not present throughout the calvarium.
2. Variation in thickness and intensity between vertices (on the same calvarium)
3. A wide variety of different calvarium types with different combinations of levels of BM adiposity, bone thickness, and subcutaneous adiposity.
4. "Blurring" between anatomic structures was present in real data and was modelled in our simulations:

a. blurring that occurred because of the voxel resolution of the MRI scanner relative to the underlying structures
b. blurring that occurred because the layers for a data point usually do not match with voxel boundaries.

The parameters used in the simulation (primarily mean and variance of both thickness and intensity of each of the different anatomical structures within a calvarium) were estimated by reviewing the MRI scans of a wide variety of real subjects from the MRI scan collections included in this study. We simulated 336 different calvaria with the following mean characteristics: 7 BM intensities (40, 50, 70, 90, 110, 140, 170) x 6 bone thicknesses (4, 6, 8, 9, 11, 12 mm) x 8 body fat thicknesses (0, 1, 2, 3, 4, 6, 9, 12 mm) and for each calvarium we generated 5000 intensity arrays. In the iterative process of simulating data, training the network, and testing its performance on real data, we observed that incorporating the characteristics of extreme head types in the simulation parameters was critical to obtaining good performance on real data. Fine-tuning of model hyperparameters played a much lesser role. The R code and all parameters are openly available [made public upon acceptance] (bmSimulation_50_versionA.r).

### Artificial neural network architecture and training

The network is a feed-forward fully connected network. There are 50 input nodes (for the 50 intensities of a datapoint), three internal layers with 100 nodes each (ReLU activation function), and six output nodes (upper and lower bound for each of outer table, BM, and inner table). The model was implemented in the Keras framework on top of the Tensorflow backend, and the training of the model was performed in Google Colab (*59*). All vertices from all calvaria were randomly shuffled, and then two thirds of the data were used for training the model, and one third was used as the validation set, to measure the performance. The loss function, defined as the sum of the mean squared errors of the six outputs, was minimised using the Adam algorithm (*60*), with a learning rate of 0.1 and a batch size of 8192. We used dropout to regularise the model (*61*), with a 30 % dropout rate in all the hidden layers. The model is trained over 300 epochs. The performance of the network plateaus before 300 epochs (Figure S1), suggesting that the optimisation has converged on the best parameters for this model architecture.

To compute the overlap of the true and the predicted intervals, the predictions were first rounded to integers, so they refer to specific layers in the intensity vector. Then the overlap is defined as the number of layers that are included in both the predicted and the true interval, divided by the number of layers in the longest of the true and the predicted interval. For each datapoint of a scan, we computed the mean BM intensity from the part of the intensity array predicted to be BM and then computed the mean and standard deviation across all datapoints in the scan. We did the same for the outer and inner table. The Python code is openly available *[made public upon acceptance]* (bmDetectNN50_randomAux_training_A.ipynb)

### Additional validation of the BMA estimation approach

In the BMA estimation pipeline, we used intensity normalized T1 images generated in the standard recon-all processing stream of FreeSurfer. To assess the effect of intensity normalization, we estimated BMA with five additional frequently used intensity normalization methods (*62*). We calculated test-retest reliability (Figure S9) and, in a dataset where quantitative T1 data was available (*57*), compared BMA estimates derived from quantitative T1 with our semi-quantitative approach, as well as each of the five additional intensity normalization methods (Figure S10). Finally, we further assessed the semi-quantitative nature of our approach by comparing outer-bone intensity with bone mineral density as measured by DEXA scans (Figure S11).

### ANN Quality control measures

We performed the BM location procedure on the intensity data from 42,068 heads in the UK Biobank. 420 of these produced no output and 2,234 produced a calvarium with less than 5,000 vertices most likely related to poor image quality (Figure S5), leaving 39,414 samples.

We used two additional QC metrics to filter out calvaria where BM location was likely to have failed. First, we set an upper limit of 30 on the standard deviation of the intensity of the outer table as scans with higher values were clear outliers and were probably cases where the location of both outer table and BM has failed (Figure S6). Second, for each calvarium, we computed the Mahalonobis distance for all vertices in the two dimensions “first layer of the BM” and “BM intensity” (Figure S7). By manual inspection we found that data points with MD > 25 often had errors in BM layer identification, typically where the network had erroneously predicted a higher and more intense layer to be the BM. We considered a calvarium as failing this QC criterium if more than 0.5% of vertices have MD > 25. This criterium is very strict as errors on only 0.5% of data points in a calvarium would not significantly affect the average BM intensity for a calvarium. However, we wanted the data to be usable to generate individual calvarial BMA maps and were only willing to tolerate a very low percentage of data points with errors in each calvarium. These two additional QC filters were complementary (Figure S8) and resulted in a final number of 39,207 scans passing QC.

### Twin heritability estimates

We used the monozygotic and same sex dizygotic twins from the *Human Connectome Project* dataset to fit the NACE model using the twinlm function of the mets package (*63*) in R (Table S1).

### GWAS methods

We followed standard quality control procedures for preparing UK Biobank genetic data. Specifically, we removed individuals missing more than 1% of genotyping data, and single nucleotide polymorphisms (SNP) missing in more than 5% of individuals or with a minor allele frequency below 0.01, yielding approximately 7.9M SNPs. We split the available sample into a white British discovery set and a non-white replication set, as detailed under *cohorts*. Within each of these samples, we performed sex-specific covariate regression prior to genome-wide-association analysis (GWAS). For males, we residualized our estimates of BM adiposity from age (using orthogonal polynomials of degree 2), scanning site, and the first 20 genetic principal components for population level stratification. For females, we applied the same model yet also controlling for hormonal replacement therapy as this is known to affect BMA (*64*) (binary: ever used HRT therapy yes/no).

The residuals were fed into GWAS using plink (version 2) (*65*). Using the discovery sample, we first performed a GWAS in females only (N=16,902), and a GWAS in males only (N=16,140). After validating their significant genetic correlation (Rg=.94, P=6e-27), we performed a GWAS in the full discovery sample, merging the residuals from the female model with the residuals from the male model. Likewise, we performed a GWAS in the replication sample merging the residuals from a male and a female model in a joint GWAS (no sex-specific GWAS performed due to power in this sample). The resulting summary statistics were standardized using the python_convert toolbox (https://github.com/precimed/python_convert), removing the MHC region in chromosome 6 between base pairs 26M and 34M before feeding them into genetic correlation analysis.

GWAS summary statistics are available on the FUMA website (ID XXXX) *[made public upon acceptance]*

GWAS results were post-processed using the Functional Mapping and Annotation of Genome-Wide Association Studies (FUMA) tool (*66*) to define genomic loci, significant independent SNPs, lead SNPs, and top lead SNPs. We used default settings which were: N = NA, Ncol = N, exMHC = 1, MHCopt = annot, extMHC = NA, ensembl = v92, genetype = protein_coding, leadP = 5e-8, gwasP = 0.05, r2 = 0.6, r2_2 = 0.1, refpanel = 1KG/Phase3, pop = EUR, MAF = 0, refSNPs = 1, mergeDist = 250. FUMA was also used to perform an eQTL analysis of the GWAS results using the GTEX v8 expression dataset. Genes, which were not the closest gene to the top lead SNP of a locus, but for which genome-wide significant SNPs were strong eQTLs, were annotated in the Manhattan plot (Figure 3).

### Single-cell data and analysis

Single cell RNAseq data of BM mesenchymal lineage cells (*30*) was downloaded from the Single Cell Portal (https://singlecell.broadinstitute.org/single_cell/study/SCP1017) and used to create a Seurat 4.0 object in R-studio (count, embedding and meta-data). This was an integrated dataset of 7585 cells that contains endosteal Td+ BM cells from 1-month-old (1M) and 1.5-month-old (1.5M) male Col2:Td mice. We did not modify the cell clustering or nomenclature, other than excluding two chondrocyte clusters as chondrocytes originate from the resting zone progenitors, but not BM MSCs (*30*). The dataset does not include lipid-laden adipocytes presumably due to the difficulty in analyzing these large and fatty cells using this method.

We searched the data from Zhong et al for the 54 genes displayed in the Manhattan plot (Figure 3). Human gene names were translated to mouse gene names and, if no similar name was identified, we searched for orthologs. For 3 of the 54 human genes, we could not identify a mouse gene and, for an additional 9 genes, the mouse gene had no expression in the scRNAseq data (Tables S7 and S8). Cluster gene expression was plotted as a dotplot (ggplot2) and patterns in gene expression were grouped by alignment to a dendrogram (ggtree) based on Euclidean distance. The expression of each gene was scaled by the gene’s mean expression across clusters (i.e. cell types). Zero expression dots were removed.

### GWAS downstream analysis and genetic architecture

We estimated SNP heritability and genomic inflation of the resulting GWAS summary statistics using LD-score regression (*67*). The same tool was used for estimating genetic correlations with bone mineral density (*28*), body mass index (*68*), waist-to-hip ratio (*69*), blood pressure (*70*) (systolic and diastolic), coronary artery disease (*71*), type-2 diabetes (*72*), multiple sclerosis (*73*), Parkinson’s disease (*74*), Alzheimer’s disease(*75*), general cognitive ability (*37*), educational attainment (*76*), and insomnia (*77*).

To estimate genetic overlap between pairs of traits we used bivariate MiXeR v1.3 (https://github.com/precimed/mixer). MiXeR(*31*) uses a Gaussian causal mixture model to estimate the total number of trait-influencing variants that explains 90% of SNP heritability (i.e., polygenicity). In this case, a trait-influencing variant is a common variant with a direct effect on the trait of interest excluding effects due to LD. Parameters are estimated with 20 iterations of the mixture models and the mean of each estimate is quantified along with the standard deviation over the 20 iterations. Given two traits of interest, MiXeR models the trait-influencing variants unique to each trait (non-overlapping) as well the number of shared trait-influencing variants (overlapping). To assess model fit, MiXeR employs the difference in Akaike information criterion between the MiXeR model and an infinitesimal model with a positive value indicative of good model fit.

### Statistical analysis

All statistical analyses are described in the relevant subsections above.

## Funding

We acknowledge the following funding sources. TK: RCN (276082, 323961). TH: RCN (295679). OAA: RCN (223273) and EU’s H2020 RIA (grant 847776). LTW: EU H2020 ERC StG (Grant 802998); RCN (223273, 249795, 248238, 286838); the South-Eastern Norway Regional Health Authority (2014097, 2015044, 2015073, 2016083, 2018037, 2018076. 2019101). We would like to thank everyone involved with the collection and sharing of imaging and genetics data used in this study.

Analysis was performed on the TSD computer facilities, owned by the University of Oslo, operated and developed by the TSD service group at the University of Oslo, IT-Department (USIT) and on resources provided by UNINETT Sigma2 - the National Infrastructure for High Performance Computing and Data Storage in Norway.

## Author contributions

Conceptualisation: TK SD TH

Methodology: TK SN TH

Investigation: TK PMB MF SN KOC TH

Visualization: TK MF TH

Writing – original draft: TK PMB MF SN KOC TH

Writing – review and editing: TK PMB MF SN KOC AF OAA LTW SD TH

## Data availability

All data are available in the main text or the supplementary materials. This research has been conducted using the *UK Biobank Resource* (*78*) (access code 27412), the *Human Connectome Project* (*58*), data from Hangzhou Normal University as part of the data provided by the *Consortium for Reliability and Reproducibility* (*25*)*, and the MICA-MICs dataset which is available on the Canadian Open Neuroscience Platform’s data portal. Human Connectome Project* data was provided by the WU-Minn Consortium (Principal Investigators: David Van Essen and Kamil Ugurbil; 1U54MH091657) funded by the 16 NIH Institutes and Centers that support the NIH Blueprint for Neuroscience Research; and by the McDonnell Center for Systems Neuroscience at Washington University. We obtained GWAS summary statistics from the following consortia: Genetic Factors for Osteoporosis Consortium, Genetic Investigation of ANthropometric Traits, ICBP International Consortium for Blood Pressure, Diabetes Genetics Replication And Meta-analysis Consortium, Coronary Artery Disease Genome-wide Replication and Meta-analysis Consortium, Complex Traits Genetics Consortium, Social Science Genetic Association Consortium, International Multiple Sclerosis Genetics Consortium, International Parkinson Disease Genomics Consortium, Psychiatric Genomics Consortium. BMA GWAS summary statistics are available on the FUMA website (ID XXXX) *[made public upon acceptance]*

## Code availability

All code and software needed to generate the results is available as part of public resources: LD score regression (https://github.com/bulik/ldsc), FUMA (https://fuma.ctglab.nl/), MiXeR (https://github.com/precimed/mixer), BM location (simulation, training, and model) *[made public upon acceptance]*.

