## Supplementary materials for "Estimating bone marrow adiposity from head MRI and identifying its genetic architecture"

Tobias Kaufmann et al.

This file includes:

Supplementary text

Figs S1 to S13

Tables S1 to S9

**Neural network training and validation on simulated data**

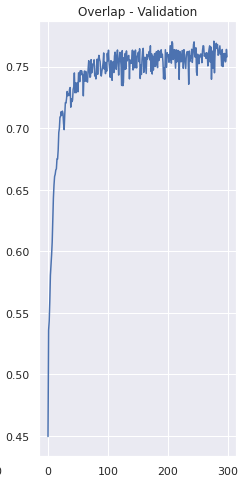

**Figure S1: Convergence of the artificial neural network before 300 epochs**

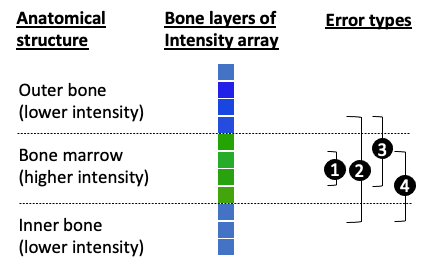

**Figure S2:** **Types of errors in BM localisation for a datapoint**

The bone marrow is typically more intense than the neighbouring bone, so only errors of type 1 usually result in overestimates of BM intensity for the given datapoint. All other error types (2, 3, 4) typically result in an underestimate of BM intensity as they incorporate less intense bone layers. This explains why the BM intensity ratio (predicted / true) averaged over all datapoints of a calvarium is centred on 0.95 and not 1 (see also S3).

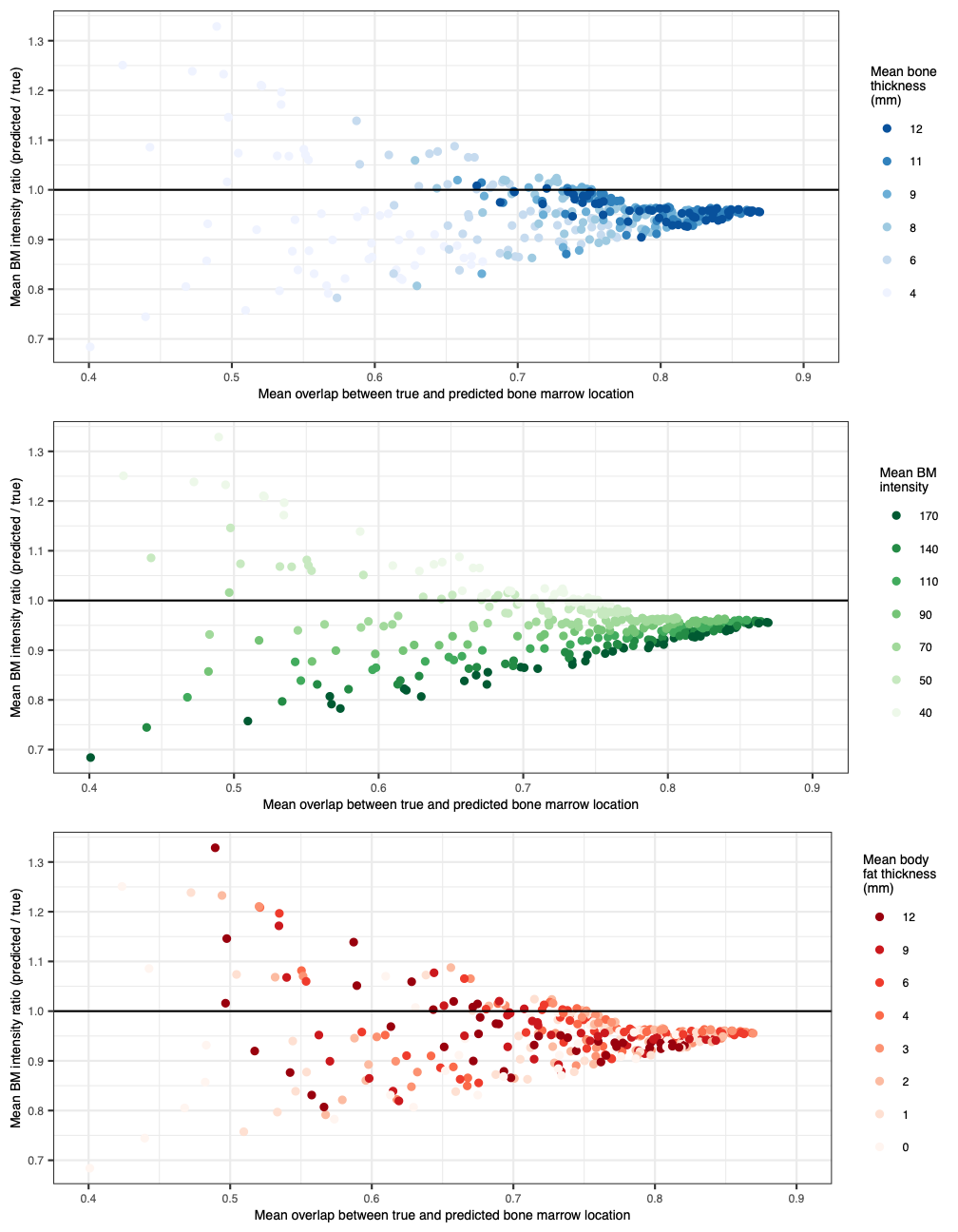
**Figure S3:** **Model performance on 336 simulated head types**

6 bone thicknesses, 7 BMA intensities, 8 body fat thicknesses).

Top panel: heads coloured by mean bone thickness.

Middle panel: heads coloured by BM intensities.

Bottom panel: heads coloured by body fat thickness.

**Tool workflow for calvarial bone marrow detection**

Main steps:

1. **Identify skin surface** using watershed <https://surfer.nmr.mgh.harvard.edu/fswiki/mri_watershed>.
2. **Identify calvarium** based on brain topology: compute mask in Matlab by reading in the parcellation from freesurfer (“aseg” file) and identifying the relevant region borders for masking the image.
3. **Extract intensity array** for every calvarium datapoint: intensity values are extracted from nu.mgz in a stepwise run from the outer skin surface inwards using mri_vol2surf from Freesurfer
4. **Localise bone marrow** for each datapoint by applying the artificial neural network model to the datapoint’s intensity array (Run_predBM in python).
5. **Average intensities located in bone marrow** for every datapoint (computeAuxiliaryFiles in R).
6. **Plot average BMA on calvarial skin surface** using Matlab (own code which draws the BMA values on the surface mesh of the outer skin surface).
7. **Average BM intensities** over all calvarium datapoints.

Software versions:

- Matlab: 2018a
- Freesurfer: 5.3.0
- R: 3.5.0
- Python: Anaconda3/5.3.0 , python 3.8.1, tensorflow 2.4.1

**Examples of BM detection from the UKB**

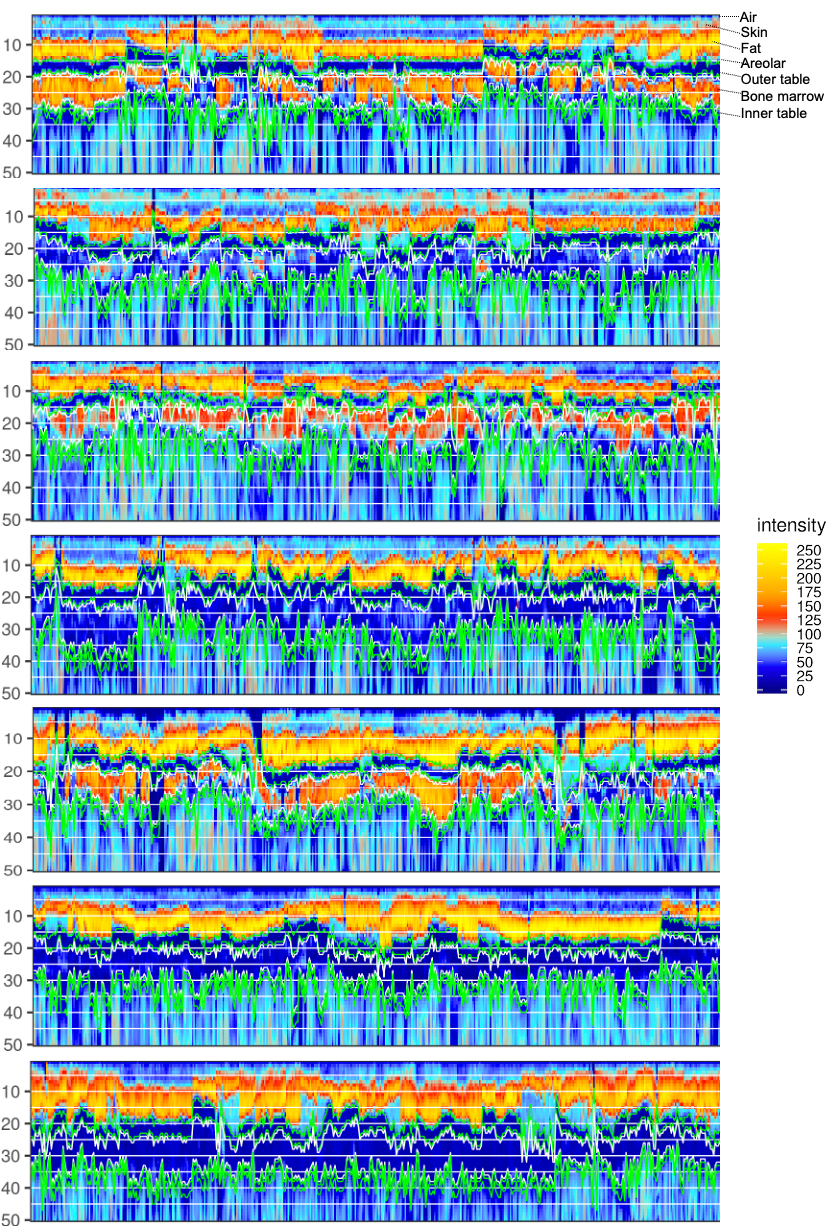

**Figure S4:** **Examples of neural network performance on seven different UK Biobank scans with different levels of subcutaneous fat thickness, bone marrow adiposity, and bone thickness**

Outer table (OT), bone marrow (BM), inner table (IT). Predicted boundaries of BM (white lines) and of outer and inner table (green lines). Adjacent datapoints (x-axis) in this plot are not necessarily adjacent on the calvarium.

**Quality control of the bone marrow localisation procedure in UKB**

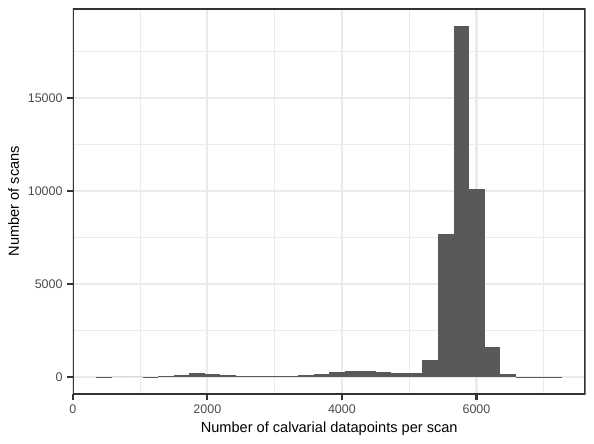

**Figure S5: Distribution of number of datapoints per scan (for 41,648 scans)**

Scans with fewer than 5000 datapoints are likely affected by poor image quality and cannot be used to reliably measure bone marrow intensity.

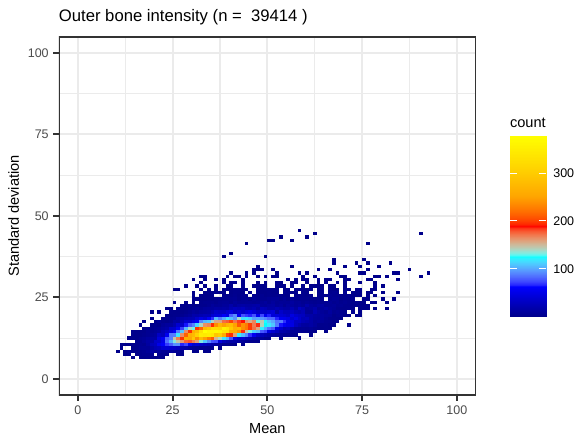

**Figure S6: Outer bone intensity mean and standard deviation for 39,414 scans**

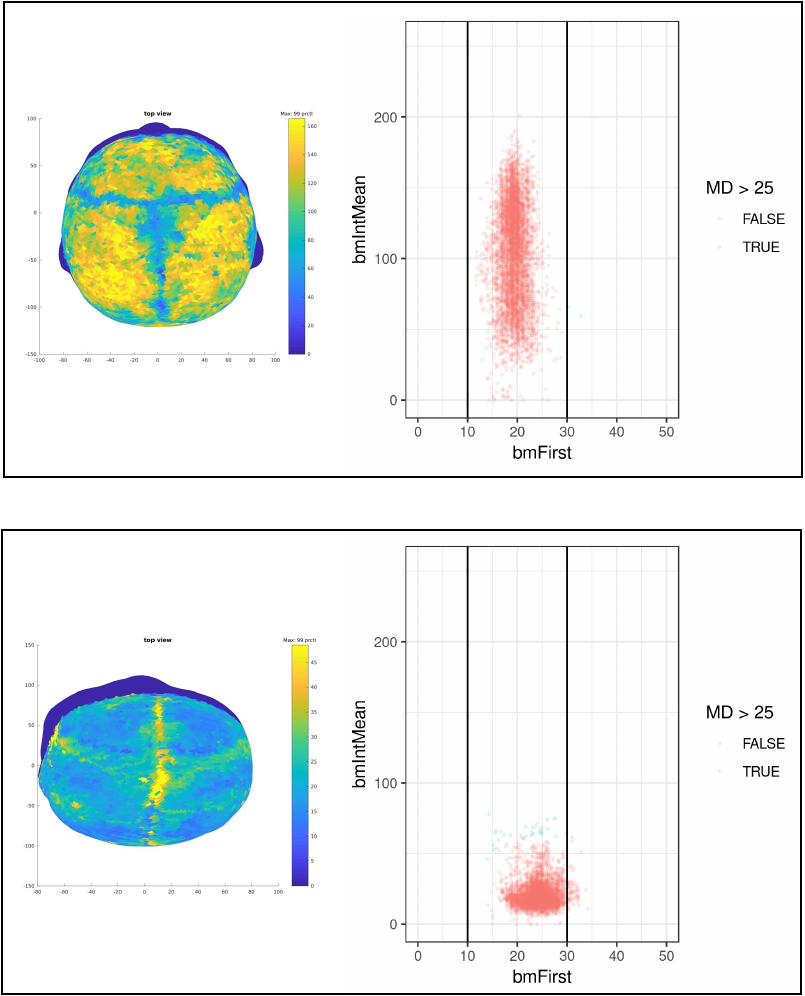

**Figure S7: BMA map of the calvarium**

BMA map of the calvarium with nose pointing north (left) and first layer of bone marrow plotted against bone marrow intensity for all datapoints in the same scan (right). Top figure for a calvarium with high levels of adiposity, but where no datapoints have a Mahalanobis distance greater than 25. Bottom figure for a calvarium with relatively low levels of adiposity, but where some datapoints have a Mahalanobis distance greater than 25. In the corresponding BMA map one can observe that these datapoints with higher intensity are all located on the sagittal suture (which is not high intensity) and correspond to datapoints in which the NN has failed to correctly localise the bone marrow layers.

 
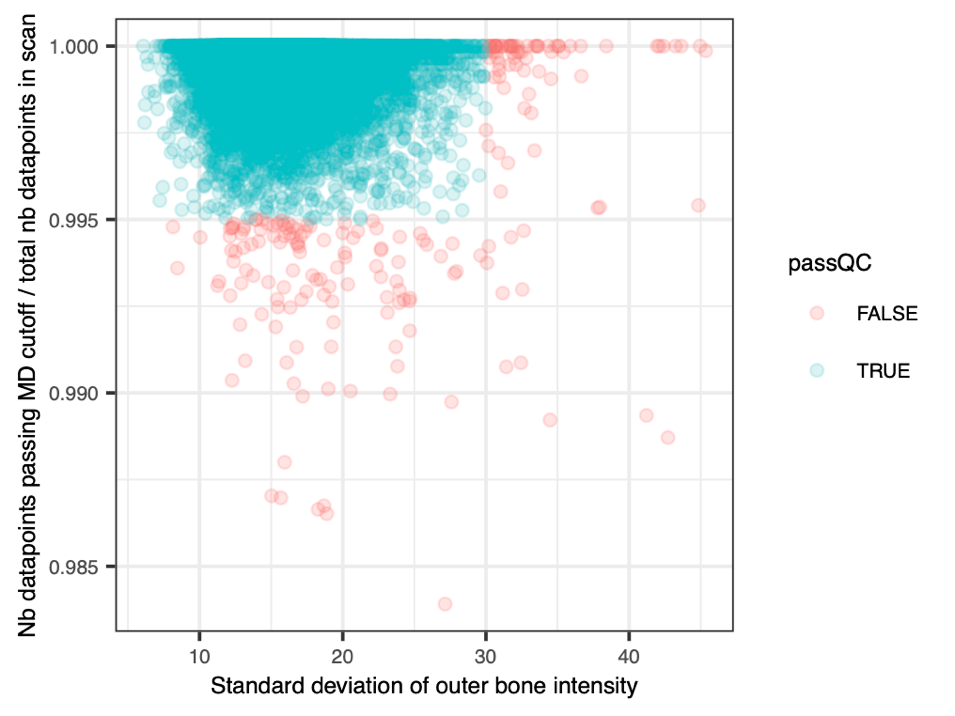

**Figure S8: Additional quality control filters**

A combination of a maximal value for the standard deviation of outer bone intensity and of a minimum value for the fraction of datapoints passing the Mahalanobis distance cutoff (see previous figure) enables additional QC of correct bone marrow localisation.

**T1-weighted MRI scan intensity as a measure of tissue composition**

*1. Comparison of different normalization approaches (HNU test-retest data):*

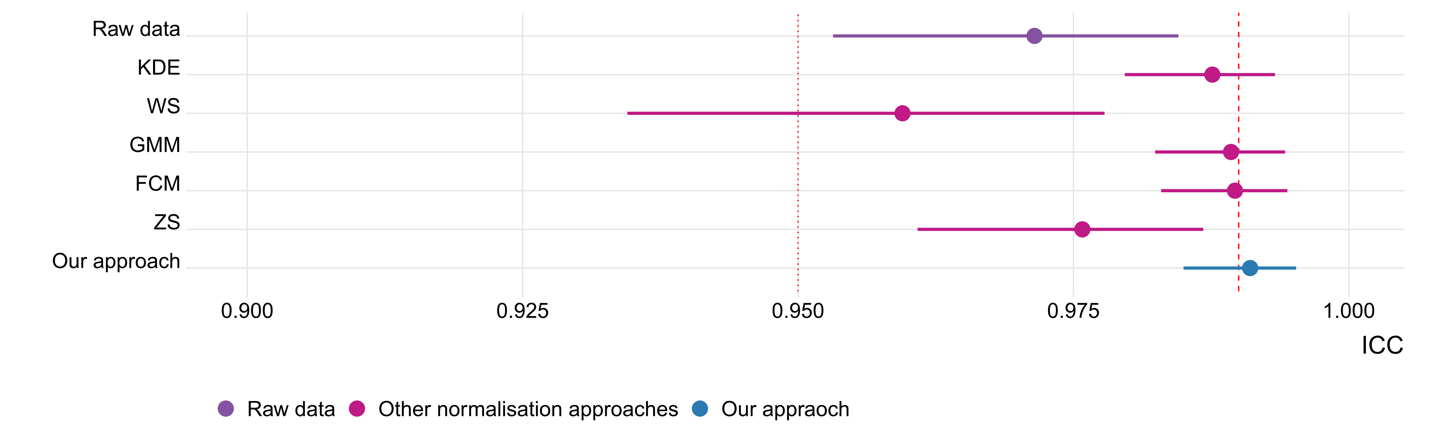
**Figure S9:** Impact of intensity normalization procedure on test-retest validity. In our proposed pipeline, T1 image intensities were rescaled to assume values between 0 and 255 as part of the recon-all processing stream of FreeSurfer. We compared this normalization approach to five additional normalization procedures that have been extensively used in the image processing literature. Specifically, we performed normalization with Kernel Density Estimation (KDE), WhiteStripe (WS), Gaussian Mixture Model (GMM), Fuzzy C-Means (FCM), and Z-score normalization (ZS). Details on these methods and their implementation are given in Reinhold et al., (2019). Using test-retest data from the HNU sample, we derived estimates of BMA for each subject and scan (30 individuals, 10 repeat scans) based on each of the five additional normalization approaches. The comparison between normalization strategies shows that several of the normalization strategies yield strong test-retest reliability, yet our approach is on par with, or better than, the other normalization strategies.

*2. Validation against quantitative T1 maps (MICA-MICs cohort):*

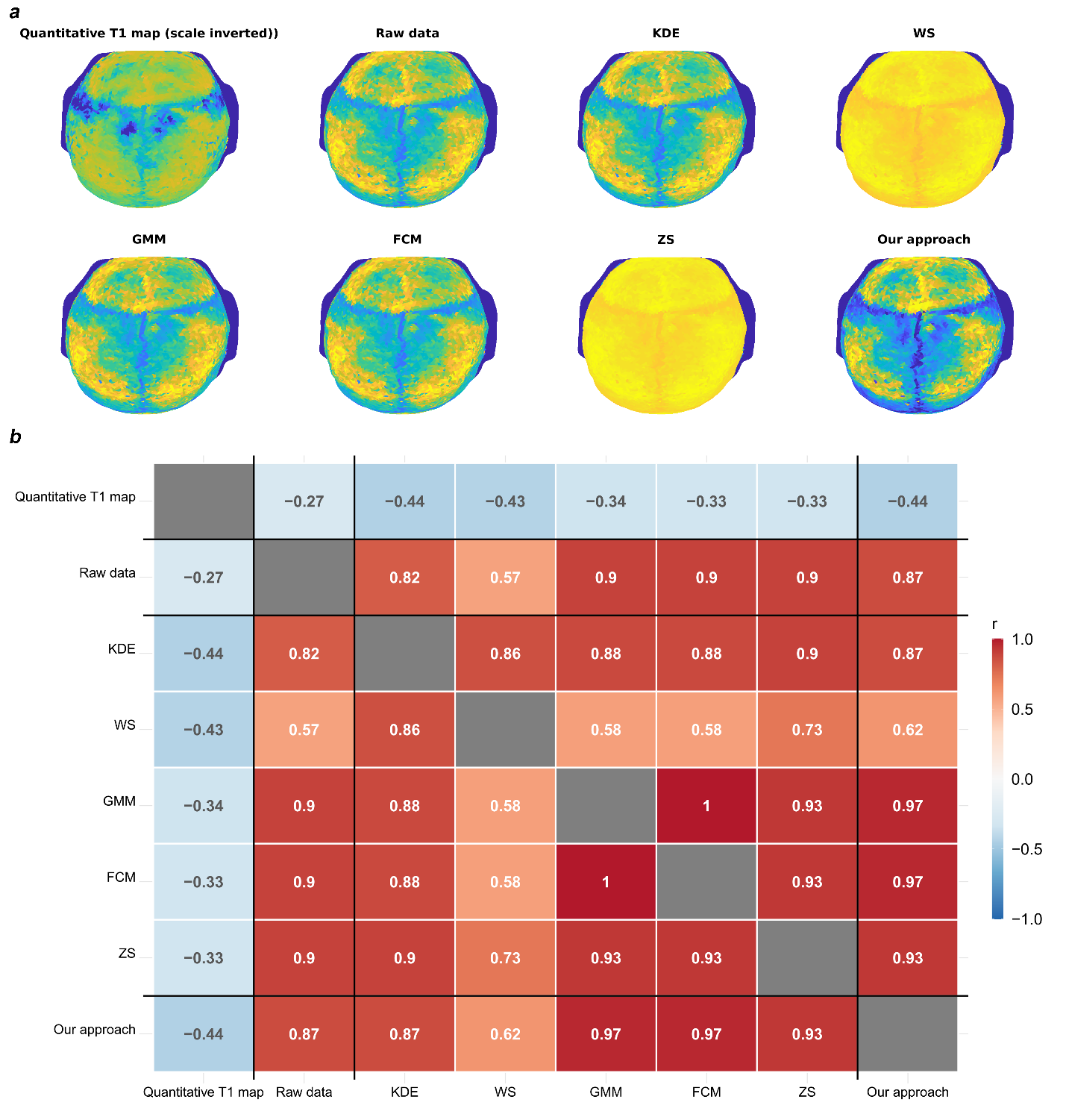

**Figure S10:** Validation against a quantitative measure of BMA**.** Data from the MICA-MICs cohort allowed us to compare our BMA estimates to BMA estimates derived from microstructurally-sensitive quantitative T1 images acquired with a MP2RAGE MRI sequence. We derived quantitative estimates of BMA from the quantitative T1 images, as well as semi-quantitative estimates based on T1-weighted images after FreeSurfer normalization (our approach) and five additional normalization approaches (KDE, WS, GMM, FCM, ZS; see also figure S9). Panel **a** illustrates the resulting calvarial maps for an example subject, and panel **b** depicts the statistical results. Our BMA estimation approach shows high correlations with estimates derived from T1-weighted images normalized with KDE, GMM, FCM, and ZS, whereas correlations between WS normalization and the other methods are lower. Most importantly, we find significant correlations between the original BMA estimation approach and fully quantitative BMA estimates. The negative correlation here is expected since T1 relaxation times are inversely correlated with fat. These significant correlations corroborate the semi-quantitative nature of our approach. We note that the correlation between quantitative and semi-quantitative approaches, while significant (p=.0017), seems moderate (our approach and KDE normalization performing best with r= -.44). However, it is important to emphasize that the age distribution of the MICA-MICs sample is very narrow (29.54 ± 5.62 years) and therefore there is little age-related variance in the derived BMA estimates. We would therefore expect even higher correlations between semi-quantitative and quantitative approaches in datasets with more variation in bone marrow adiposity, such as data covering a wider age span or life events such as menopause.

*3. Correlation between outer-bone intensity and bone mineral density (UKBiobank):*

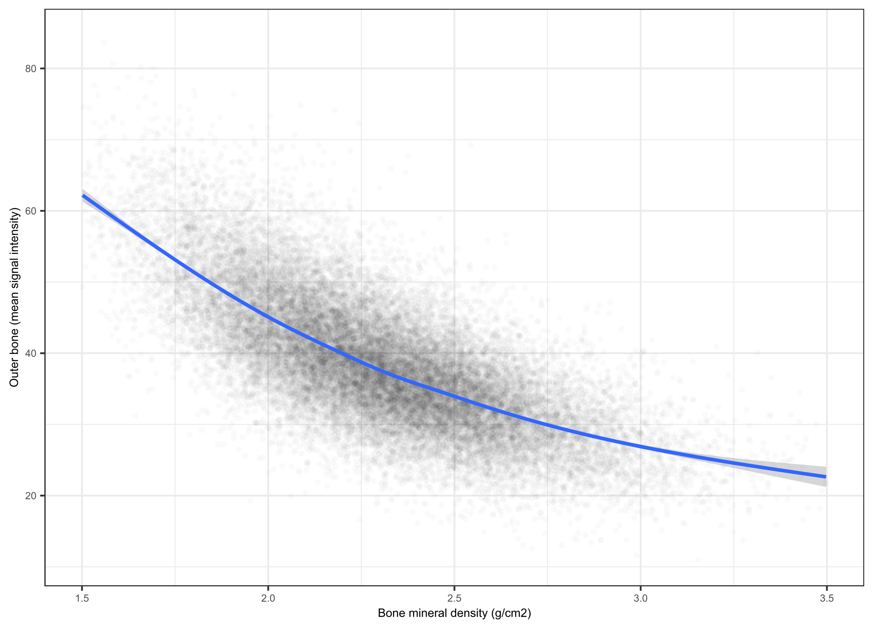

**Figure S11**. Illustration of the semi-quantitativeness of another measure obtained from T1-weighted MRI. To further illustrate that semi-quantitative approaches from T1-weighted scans can provide plausible measures, we have additionally included an analysis of another calvarial bone measure, specifically the outer-bone intensity. Bone mineral density measures the mineral content of bone in g/cm2 and correlates negatively with cortical bone signal intensity in T1-weighted images. We can therefore obtain a second independent estimate of the quantitativeness of signal intensity from the comparison of mean outer-bone signal intensity with calvarial bone mineral density (measured by DEXA scan). We estimate a correlation of -0.715 (CI [-0.72,-0.71]) between these variables in the 29,093 individuals that had both MRI scans and calvarial bone mineral density measurements. This correlation is most likely an accurate estimate of the quantitativeness of the signal intensity as there is a wide distribution of bone mineral density in the 29,093 individuals used for this estimate.

*4. Significant difference in bone marrow adiposity between patients with osteoporosis and control individuals:*

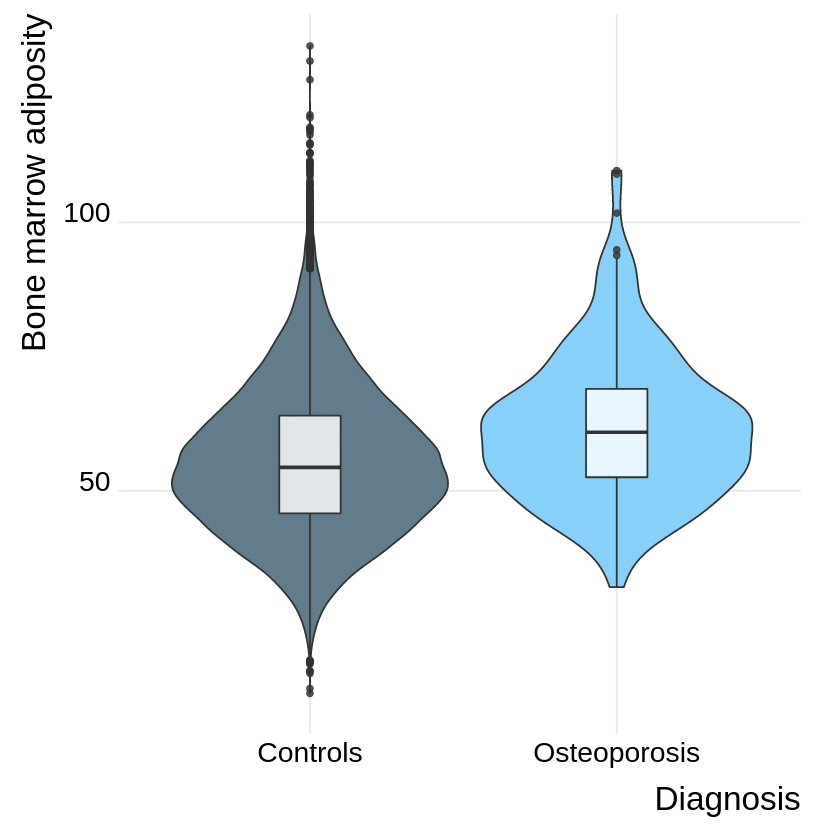

**Figure S12.** Significant difference in bone marrow adiposity between patients with osteoporosis and control individuals without a diagnosis within the ICD-10 M category.

We identified a set of N=220 individuals with an ICD-10 diagnosis of osteoporosis (ICD-10 codes M80-82) in the UK Biobank dataset and compared this group with N=30,127 controls, excluding individuals with any other diagnosis within the M category. In the models adjusted for age, sex, and scanner, BMA was significantly higher in the osteoporosis group (t=9.3; p<2e-16). Among females, the effect was similar also when controlling for HRT use (t=7.3; p=3e-13). This analysis shows that our proposed BMA measure allows for robust comparison between subjects and provides another replication of a well-established biological relationship, i.e., BMA increase in osteoporosis.

**Twin-based heritability estimates in HCP**

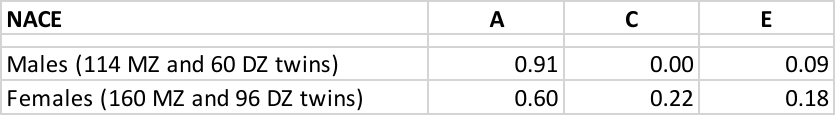

**Table S1: Twin-based heritability estimates** using the NACE model fitted in R using the twinlm() function of the mets package. NACE: normal (N) additive genetic effects (A), common environment (C), the unique environment (E) Female pairs aged 26 to 36 and male pairs aged 22 to 34. The Falconer method produced similar heritability estimates.

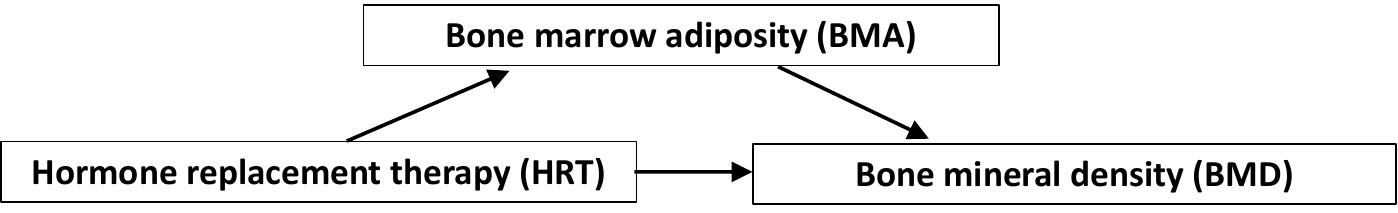

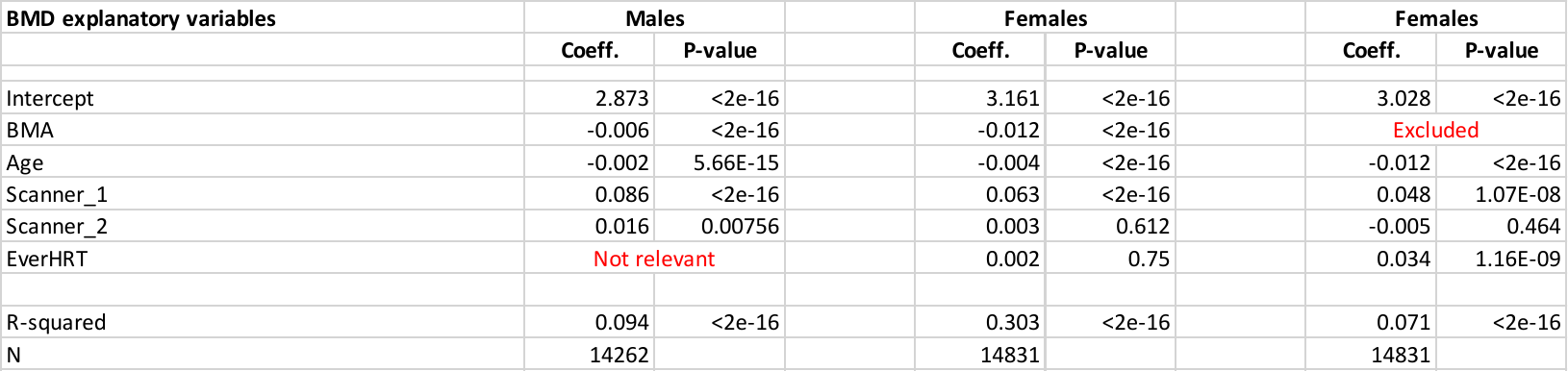

**Table S2: OLS regression analysis of the determinants of BMD (in males and females)**

Two models are shown for females, one including BMA (left) and one excluding BMA (right). Age and HRT have statistically significant effects on BMD when BMA is excluded. The coefficient on HRT drops by a factor of 20 and loses statistical significance when BMA is included in the regression and R^2^ of the regression increases by a factor of 4, thus indicating that the effect of HRT on BMD is largely mediated by HRT’s effect on BMA. The number of samples is somewhat reduced because BMD data was not available for all samples with BMA data.

**GWAS quality control and genome-wide significant loci**

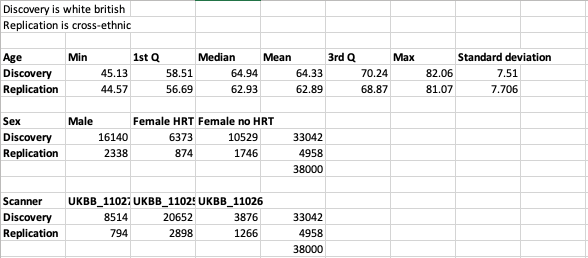

**Table S3: Characteristics of the discovery and replication samples**

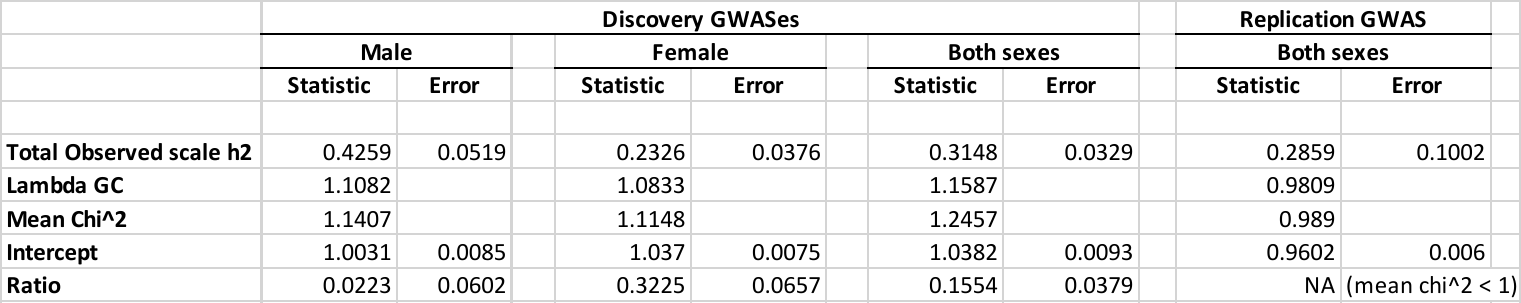

**Table S4: LDSR estimates of h^2^_SNP_ and genomic inflation**

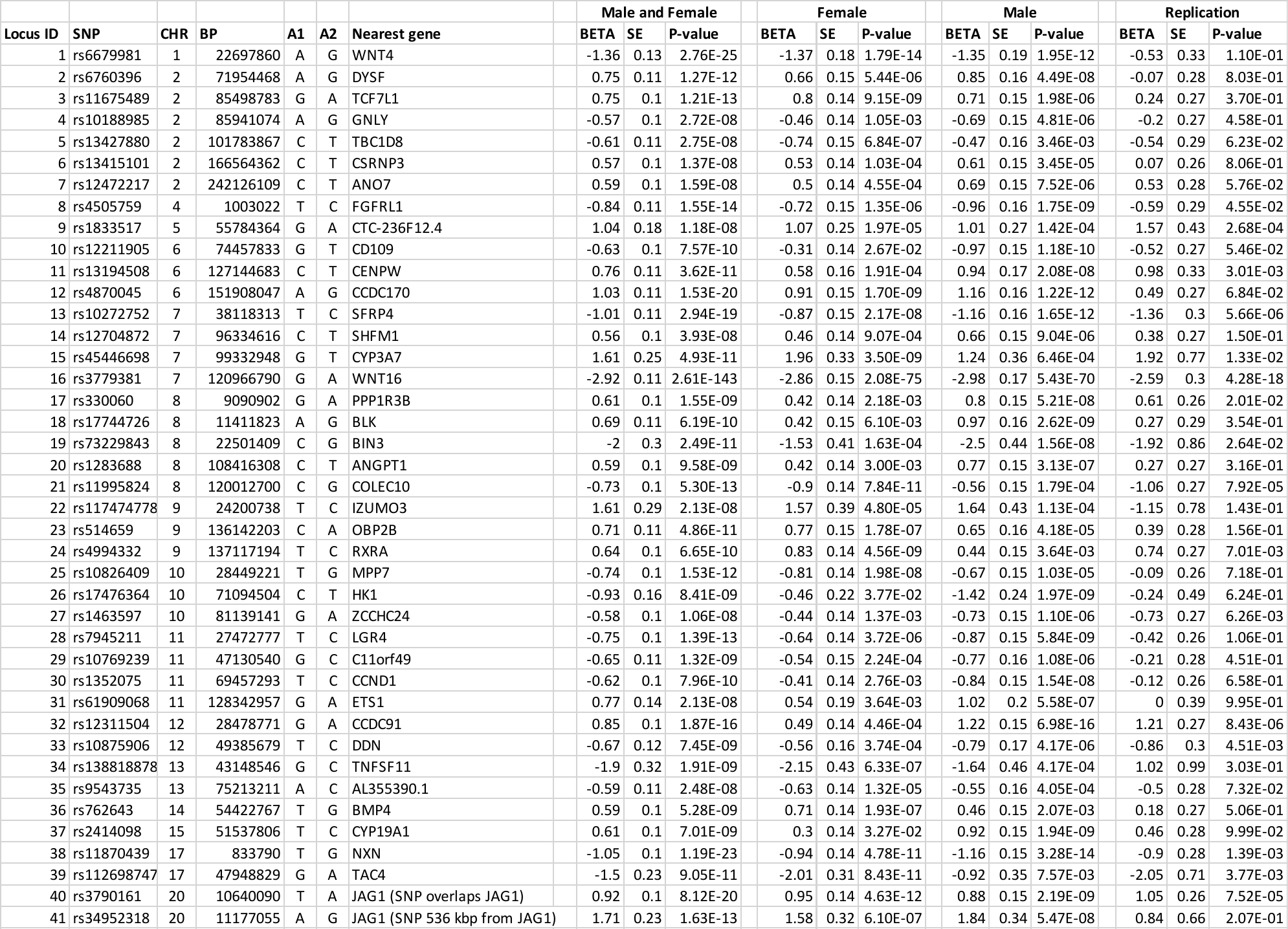

**Table S5: Discovery genome-wide significant loci (male, female, and both sexes) and replication (both sexes)**

92.7% of the lead SNPs of the discovery sample showed same effect direction in the replication sample.

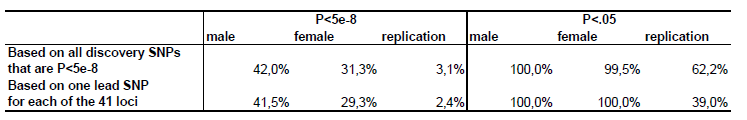

**Table S6: Comparison of p-values between the “both sexes” discovery sample and the male, female, and replication (both sexes) samples.**

**FUMA results**

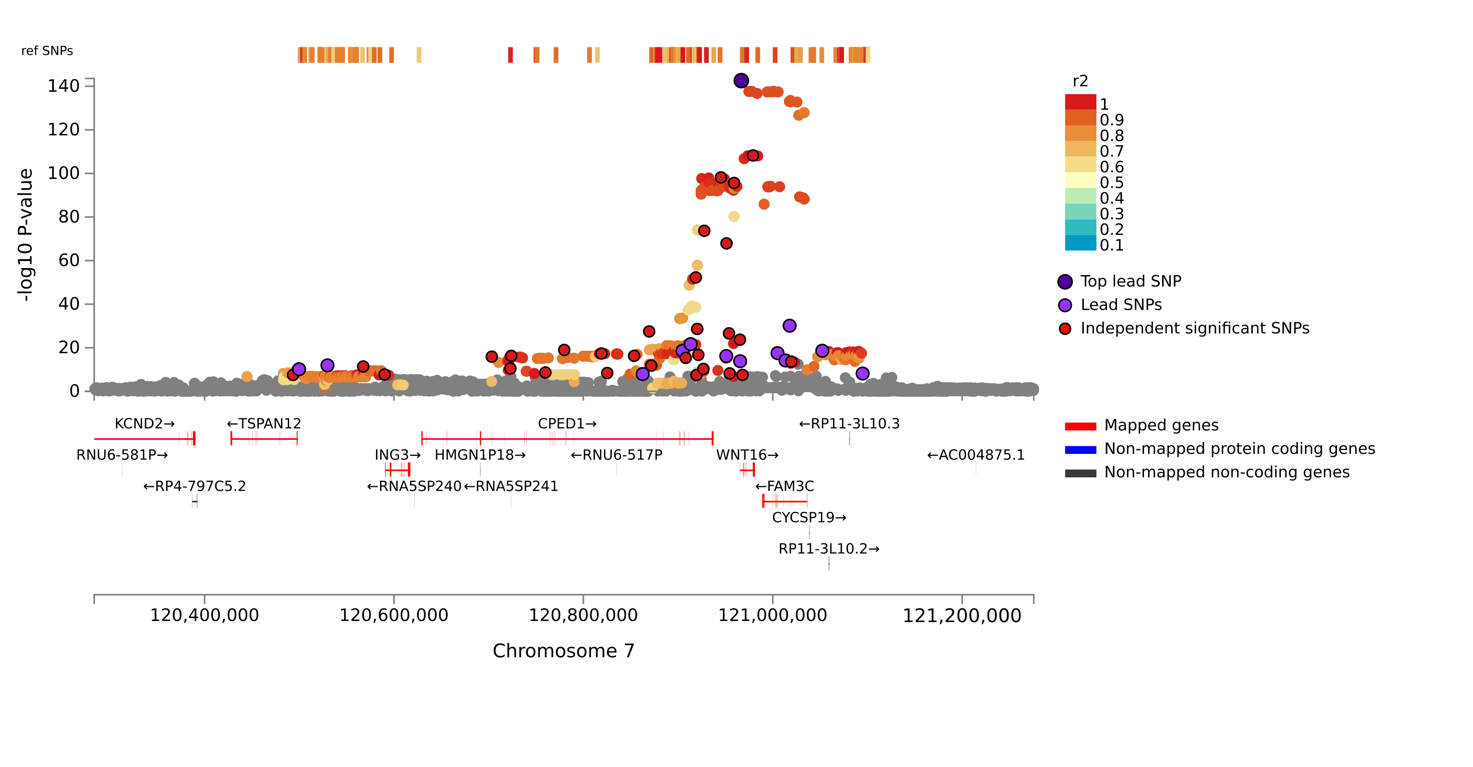

**Figure S12 The WNT16 locus in the joint discovery sample**

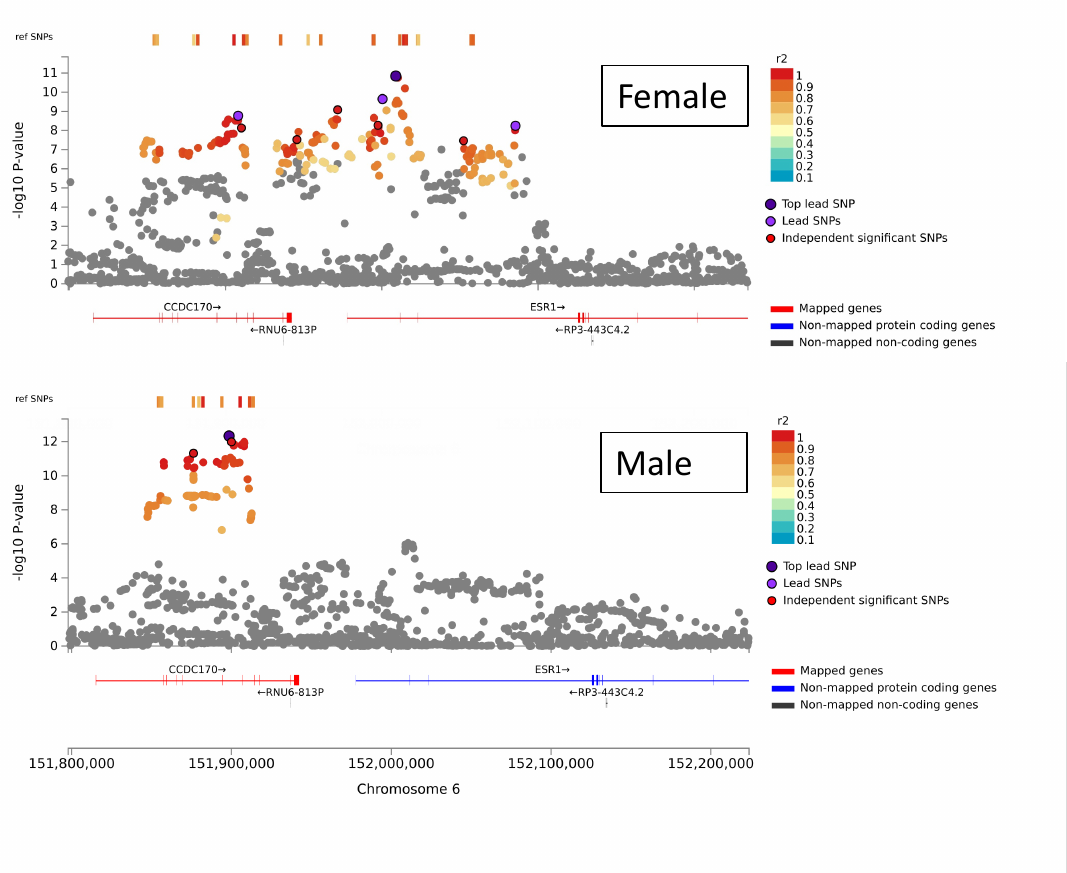

**Figure S13: The ESR1 locus in the male and female discovery samples**

**Single-cell RNAseq data from mouse bone marrow**

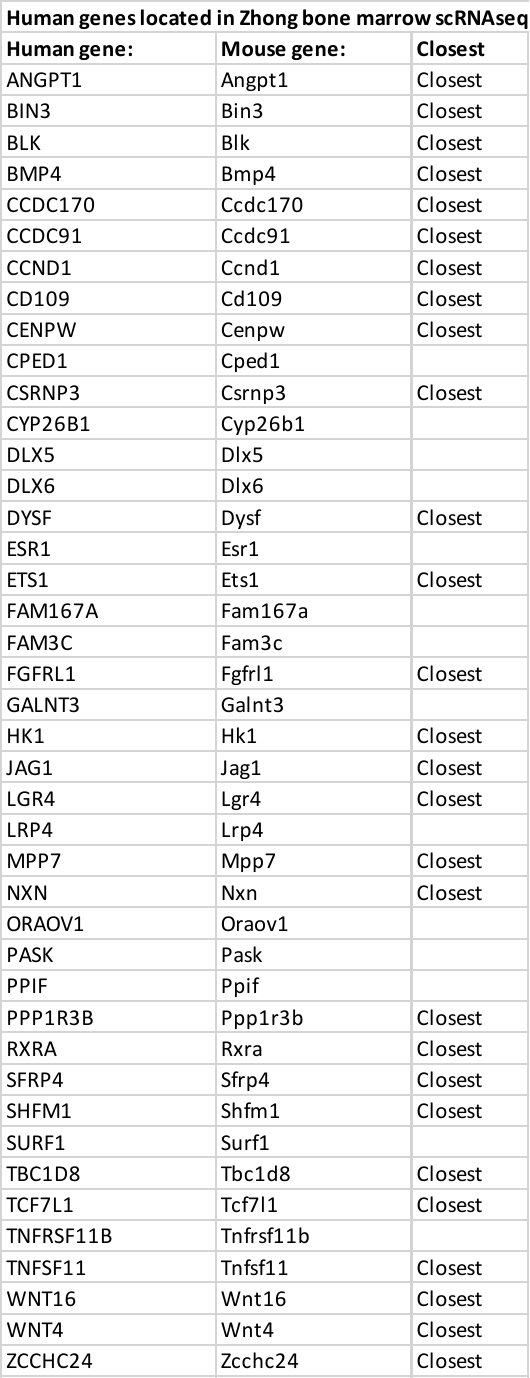

**Table S7: Human genes for which expression data was located in the Zhong dataset**

Closest indicates that the gene is closest to the lead SNP in a locus. Genes in the table not marked closest are genes for which SNPs in the locus are strong eQTLs.

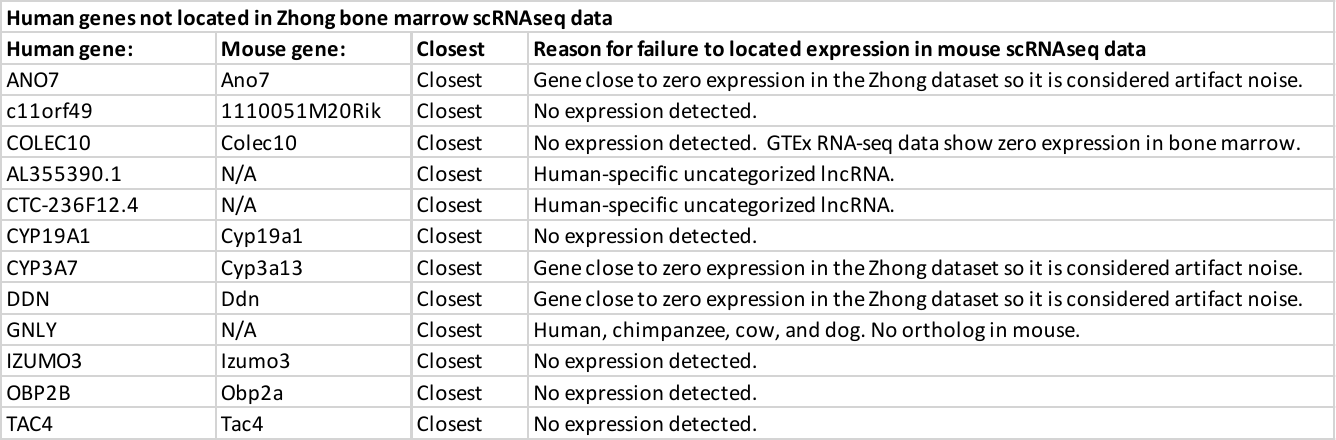

**Table S8: Human genes for which expression data was not located in the Zhong dataset**

**Genetic overlap between BMA and other traits at the locus level**

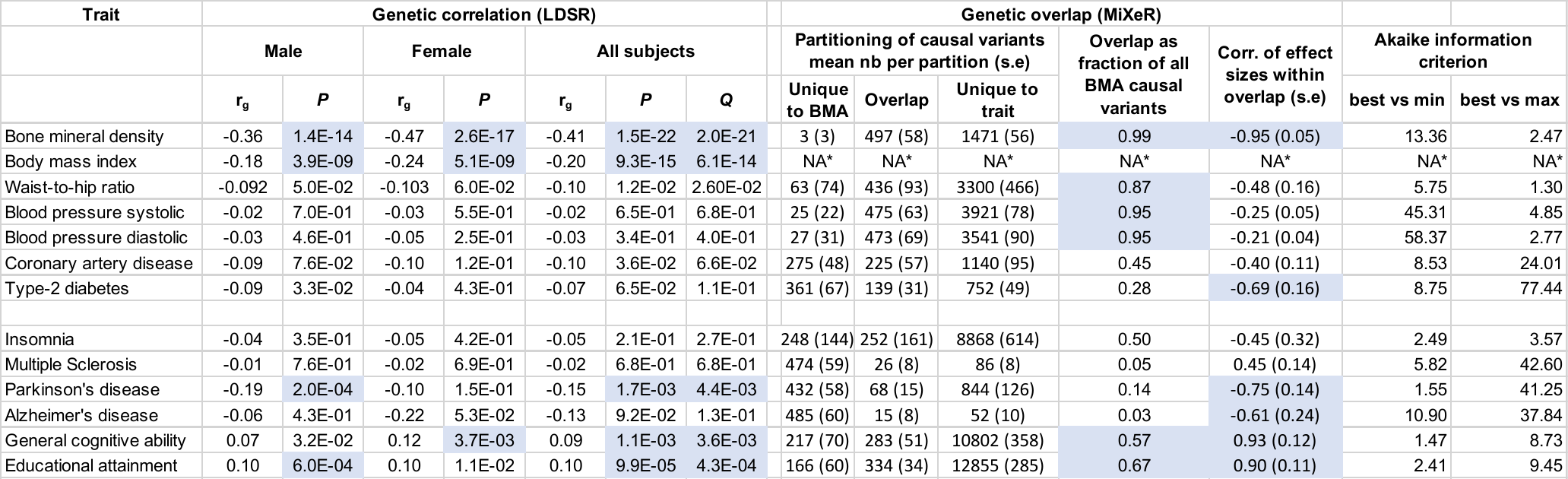

**Table S9: Akaike information criterion values for all MiXeR models**

Positive best_vs_min_overlap AIC estimates indicate that MiXeR can accurately distinguish the reported overlap from the minimum possible overlap allowed.

Positive best_vs_max_overlap AIC estimates indicate that MiXeR can accurately distinguish the reported overlap from the maximum possible overlap allowed.
